# Simultaneous assessment of genetic and epigenetic contributions to plasma lipid levels with respect to cardiovascular risk

**DOI:** 10.1101/2024.05.21.24307654

**Authors:** Fumihiko Takeuchi, Masaya Yamamoto, Masahiro Nakatochi, Kozue Takano, Atsuko Okazaki, Sakurako Emoto, Yasuharu Tabara, Tomohiro Katsuya, Ken Yamamoto, Masato Isono, Kotaro Mori, Tatsuaki Matsubara, Sahoko Ichihara, Mitsuhiro Yokota, Hisao Hara, Yukio Hiroi, Norihiro Kato

## Abstract

**Background:** This study aims to develop a model for simultaneously assessing genetic and epigenetic contributions to plasma lipid levels.

**Methods:** The predictive model was developed using two cardiovascular risk groups, i.e., individuals with high low-density lipoprotein cholesterol (LDL-C) levels (≥160 mg/dl, *N* = 296) and coronary artery disease (CAD) (*N* = 315), in contrast to reference (max *N* = 3,801) and non-CAD individuals (*N* = 164). For genetic predisposition, rare pathological variants in five target genes related to familial hypercholesterolemia (FH) were screened, while common variants were characterized to calculate a polygenic risk score (PRS). The methylation risk score (MRS) was also calculated for epigenetic profiles based on DNA methylation levels at 13 CpG sites. A relationship between these variables and lipid levels was analyzed in regression and quantile models.

**Results:** A total of 17 rare FH-related gene variants were identified in patients with high LDL-C or CAD, significantly more prevalent than in the general Japanese population (2.8% vs. 0.2%, *P* <1×10^−15^). For the rare variants plus PRS, the predictability of individual LDL-C increased (correlation coefficient between predicted and measured values, *r* = 0.261, *P* = 1.7×10^−11^) compared to PRS alone (*r* = 0.151, *P* = 1.2×10^−4^). PRS and MRS had the most significant impact on high-density lipoprotein cholesterol and triglycerides, respectively. The two risk scores had additive effects on these traits.

**Conclusions:** Our results provide proof-of-concept that assessing the relative contribution of genetic predisposition and DNA methylation levels (reflecting past environmental exposures) may help individuals refine their dyslipidemia treatment.

---

Dyslipidemia is a significant risk factor for atherosclerotic cardiovascular diseases, including coronary artery disease (CAD), to which genetic, environmental, and demographic factors contribute interactively. Epidemiological studies have shown that appropriate management of dyslipidemia can significantly decrease cardiovascular morbidity and mortality. Atherosclerosis treatment has often focused on reducing low-density lipoprotein cholesterol (LDL-C) among dyslipidemias, with lifestyle modifications emphasized in all patients with high LDL-C.^1,2^ Above all, dietary intervention is essential because high saturated fat intake causes increased concentrations of LDL-C.^3^ However, sustaining lifestyle modifications over time is generally challenging, so drug therapy is sometimes necessary to reduce the risk of atherosclerosis.^1^

People with a vital genetic component, e.g., familial hypercholesterolemia (FH) patients, also require lipid-lowering medications such as statins at an earlier stage.^2^ For CAD, while the introduction of statins has considerably changed its management and prevention, the disease burden remains high. Clinical practice guidelines emphasize the importance of estimating the absolute risk of CAD and tailoring the intensity of preventive actions accordingly.^1,4^ The optimal LDL-C goal for persons at low risk is set at <116 mg/dl by the 2019 ESC Guidelines, which used to be set at <160 mg/dl (for persons with 0-1 risk factor) by the NCEP-ATPIII Guidelines. Still, there is a significant gap between guideline-recommended treatment and daily clinical practice of dyslipidemia.^5^

An individual’s lipid levels are influenced by the degree of environmental exposure and genetic predisposition, although no method has been established to provide quantitative indicators of these combinations. On the one hand, it is difficult to quantitatively assess the impact of unhealthy lifestyle habits on lipid levels. Even so, exposure to some external factors, such as smoking and high plasma lipids, can cause long-lasting DNA methylation changes in blood cells.^6–8^ These changes can reflect an individual’s environmental history, and consequently, lifestyle habits may be evaluated through epigenetic biomarkers.^9^ On the other hand, genetic information as an auxiliary diagnostic tool for dyslipidemia is still in the realm of research, with a few exceptions, such as the genetic diagnosis of FH.^2^ In this line, polygenic risk score (PRS) has recently become popular to summarize the cumulative effects of genetic loci, successfully identified by genome-wide association studies (GWAS) of lipid traits.^8,10^ Nonetheless, there remains considerable uncertainty in individual PRS estimation for PRS-based risk stratification^11^ and substantial difficulty in unbiased assessment of the contribution of rare variants to complex traits.^12^

In light of this situation, we performed genetic (i.e., GWAS array genotyping and rare variant search in FH-related target genes) and epigenetic (specifically, DNA methylation) analyses focusing on lipid traits, specifically LDL-C. We aim to develop a model for evaluating genetic and epigenetic contributions to plasma lipid levels with a risk prediction index simultaneously incorporating multiple data modalities for individuals with high LDL-C or CAD. We studied Japanese populations to overcome challenges in applying PRS and methylation profiles to non-European ancestry groups.^13,14^ We collected individual-level data to test our risk prediction index’s accuracy. The study demonstrates that a combination of genetic predisposition and DNA methylation has an additive effect on an individual’s lipid levels among people at high risk of atherosclerosis and supports the potential utility of multi-omics data in refining an individual’s dyslipidemia treatment.

## Materials and Methods

Please see the Data Supplement for details about the Materials and Methods.

### Study population

The institutional ethics review board approved this study, and participants gave their written informed consent. The procedures followed the ethical standards of the institutional committee on human experimentation at the National Center for Global Health and Medicine (NCGM).

In NCGM, adult patients of Japanese descent were recruited via two separate projects: BIO-CVD and NCGM Biobank, whose enrollment started in 2014 and 2012, respectively. They were categorized into three subgroups based on their plasma LDL-C levels and CAD status (see Supplementary Methods): high LDL-C without CAD, CAD, and non-CAD subgroups (as shown in Table 1). A hospital-based study called BIO-CVD assessed CAD in 475 participants (306 CAD and 169 non-CAD). Samples from the NCGM Biobank were also examined, specifically those with elevated plasma LDL-C (≥160 mg/dl) (*N* = 300). These samples were used for genetic and DNA methylation analyses (Fig. 1). Also, a significant number of Japanese individuals from non-targeted populations were included. We used publicly available whole genome sequencing data (*N* = 8.3K) for low-frequency (or rare) genetic variants. For common single nucleotide variants (SNVs), we used GWAS array genotyping data in three population-based cohorts (including the Kita-Nagoya Genomic Epidemiology [KING] study cohort^15^) and another hospital-based sample at NCGM (named NCGM hospital cohort; *N* = 351) (Table S1), which we referred to hereafter as reference individuals in total (*N* = 3,801). Furthermore, for DNA methylation, we utilized epigenome-wide association study (EWAS)-array assayed data for part (*N* = 314) of the KING study cohort samples previously reported.^15^

**Fig. 1.**
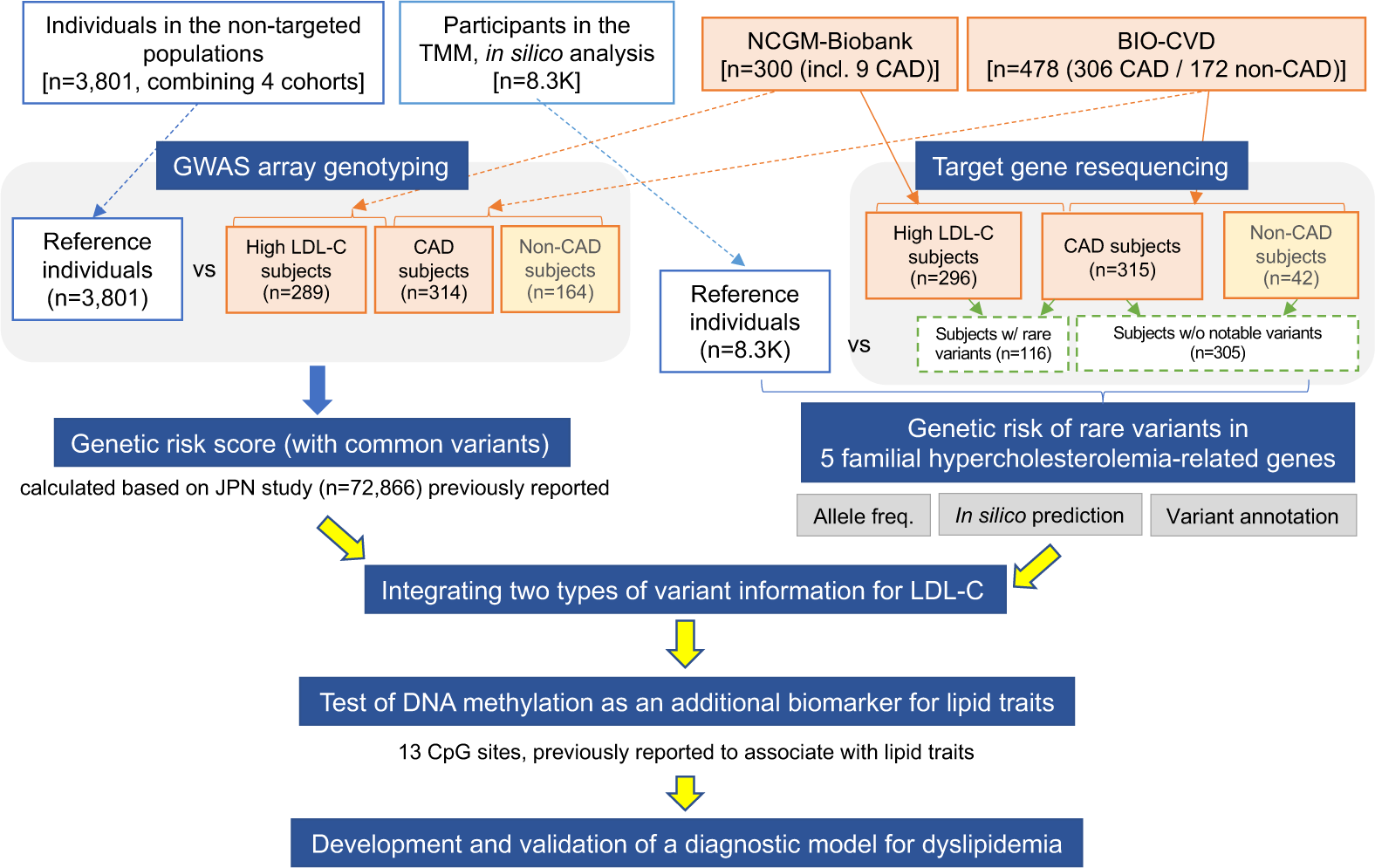
Overview of study design. The integrative genomic analysis involves GWAS array genotyping, target gene resequencing, and measuring DNA methylation at selected CpG sites. In contrast to two cardiovascular risk groups— high LDL-C and CAD subgroups, individuals in the non-targeted populations (*N* = 3,801) were used as reference datasets for GWAS array genotyping. The non-CAD subgroup (*N* = ∼164) was compared to the CAD subgroup by GWAS array genotyping and target gene resequencing. Participants in Tohoku Medical Megabank (TMM) (*N* = 8.3K) were also used as large *in silico* gene+c datasets for the evalua+on of rare variant frequencies. Moreover, part (*N* = 314) of the KING study cohort was used as a reference dataset for DNA methyla+on analysis.

**Table 1.** Participant characteristics.

| | High LDL-C ($\geq 160$)<br>subjects w/o CAD | CAD subjects | non-CAD subjects |
| --- | --- | --- | --- |
| No. of individuals (F/M) | 296 (151/145) | 315 (51/264) | 164 (54/110) |
| Age, yr | 64.0 ± 0.7 | 68.7 ± 0.6 | 69.7 ± 0.7 |
| BMI, kg/m<sup>2</sup> | 23.6 ± 0.2 | 24.5 ± 0.2 | 23.6 ± 0.3 |
| LDL-C, mg/dl | 179.4 ± 1.3 | 98.6 ± 1.7 | 109.5 ± 2.1 |
| HDL-C, mg/dl | 60.4 ± 1.1 | 48.7 ± 0.7 | 56.7 ± 1.1 |
| Triglycerides, mg/dl | 144.3 ± 4.1 | 143.8 ± 4.4 | 129.4 ± 5.5 |
| Smoking habit* |  |  |  |
| Never, n (%) | 142 (48%) | 90 (29%) | 61 (37%) |
| Former, n (%) | 113 (38%) | 153 (49%) | 76 (46%) |
| Current, n (%) | 41 (14%) | 70 (22%) | 27 (16%) |
| Complication** |  |  |  |
| Hypertension, n (%) | 153 (52%) | 245 (78%) | 114 (70%) |
| Diabetes, n (%) | 52 (18%) | 130 (41%) | 46 (28%) |
| Hyper-LDL-cholesterolemia, n (%) | 296 (100%) | 152 (48%) | 48 (29%) |
| Statin, n (%) | 57 (19%) | 221 (70%) | 47 (29%) |
| Ezetimibe, n (%) | 13 (4%) | 32 (10%) | 2 (1%) |
| CAD, n (%)*** | — | 315 (100%) | 0 (0%) |

Samples in the table were derived from either the BIO-CVD project or the NCGM Biobank and all but 122 non-CAD individuals were used for resequencing of 5 target genes related to familial hypercholesterolemia (see Methods). CAD, coronary artery disease. LDL-C, low density lipoprotein cholesterol. Of 315 CAD patients, 157 had a clear history of myocardial infarction (MI) but 158 did not: an average number of diseased vessels, 2.0 (0.1) vs 1.8 (0.1); the age of CAD onset, 63.0 yr (1.0) vs 66.9 yr (1.0); left ventricular ejection fraction in echocardiography, 50.7% (1.0) vs 63.9% (0.4) [MI vs non-MI, SE in parentheses].
\*The smoking status of 2 individuals in a CAD group is unknown.
\*\* Hypertension and diabetes are defined when individuals are prescribed anti-hypertensive and anti-diabetic drugs, respectively. Hyper-LDL-cholesterolemia is defined when basal LDL-C

When analyzing prediction models for dyslipidemia, the high LDL-C and CAD subgroups were set as atherosclerotic risk groups, while the KING study cohort samples and the non-CAD subgroup were set as comparison groups (Fig. 1). With these subgroup analyses, we attempted to test whether high LDL-C or CAD status affects the association between DNA methylation and lipid traits.

Medical records were reviewed to extract the participants’ plasma lipid levels and medication information at enrollment and clinical conditions of CAD patients (Table 1). To ensure the reliability of the data, we excluded individuals who had missing information on key variables: plasma lipid levels, genotype, and DNA methylation data.

### Imputation of baseline LDL-C level

For the imputation of baseline (or pretreatment) LDL-C, we performed a retrospective analysis on data from 36 individuals with high LDL-C in whom plasma LDL-C measurements were available both before and within 18 months after initiating statins and ezetimibe (Fig. S1). Similar to the study by Ruel *et al.,*^16^ we performed imputation of baseline LDL-C by dividing the on-treatment value by the proportion obtained by subtracting the expected treatment-induced reduction (expressed as a ratio) from 1.

### Gene sequencing

Next-generation sequencing screened mutations in five FH-related genes (*LDLR*, *APOB*, *PCSK9*, *LDLRAP1*, and *APOE*). The sequences were aligned to a human genome reference sequence (GRCh37/hg19). SNVs and insertion/deletion (Indel) were called using the software GATK (https://gatk.broadinstitute.org/) and McCortex (https://github.com/mcveanlab/mccortex) and annotated using VarSeq (https://www.goldenhelix.com/products/VarSeq/). We then retrieved rare, putatively pathogenic variants, including SNVs that cause non-synonymous, nonsense, or splice-site substitutions and are predicted to be deleterious by ClinVar, LOVD, or *in silico* prediction algorithms in dbNSFP.^17^

### Genotyping

767 samples were newly genotyped with the Infinium OmniExpress-24 BeadChip (Illumina, San Diego, CA, USA). Besides, GWAS array genotyping data for reference individuals^18^ were utilized.

### Polygenic risk score (PRS)

For each lipid trait, a PRS was built using LDpred (https://github.com/bvilhjal/ldpred)^19^ and external GWAS summary statistics,^20^ for which potential population stratification was adjusted using genetic principal components.^21^ Briefly, for each linkage disequilibrium (LD)-based clump across the genome, index SNPs passing specific statistical significance (*P* <0.005) and LD coefficient (*r*^2^ ≥0.5) thresholds were extracted from large GWAS reported in Japanese.^20^ Individuals were scored based on the number of risk alleles (0, 1, or 2) weighted by beta of the SNP-trait association, thereby used for developing the best polygenic predictive indicator of LDL-C (PRS_LDL-C_) and other lipid traits.

### Conversion of PRS to LDL-C value

We developed a formula that converts PRSs to LDL-C values using two different external reference panels to estimate the standardized effects of a PRS_LDL-C_ on plasma LDL-C level. First, we set standardized PRS_LDL-C_ decile classes in the general Japanese population and used them as the Japanese standard PRS for LDL-C (Fig. S2A-C). Then, we calculated the mean LDL-C values for Japanese standard PRS decile classes in the NCGM hospital cohort, which was not previously screened for cardiovascular disease (Table S1 and Fig. S3). The mean values of LDL-C for each PRS decile class (shown in Fig. S2D) were used to convert PRS_LDL-C_ in individuals from BIO-CVD and NCGM Biobank (Fig. 3D and Fig. S2E) belonging to the same population as the NCGM hospital cohort.

### Genetic composite risk

For rare [minor allele frequency (MAF) <0.01] variants of putative functional significance, we arbitrarily classified them into three distinct categories—(1) disruptive (frameshift or splice-donor), (2) damaging, and (3) all other non-synonymous variants—similar to the previous study^22^ with some modifications (see the Supplementary Methods). We estimated the net increase in LDL-C levels for rare variant carriers by subtracting a predicted value (based on PRS_LDL-C_) from the actual measured value. Using linear regression models, we calculated LDL-C changes from individuals without rare FH-related gene variants and the standard error (SE) by category, including disruptive, damaging, and other non-synonymous variants.

To combine the genetic risk of rare variants and SNVs, we used the following algorithm: [an individual’s LDL-C value] = [predicted LDL-C value based on PRS_LDL-C_] + [estimated changes from non-carriers (by rare variant category)].

### Methylation risk score (MRS)

Genomic DNA was extracted from the peripheral blood buffy coat and stored at −80°C for DNA methylation analysis. As previously reported,^23^ genome-wide methylation profiling was performed with the Illumina EPIC array (Illumina Inc. San Diego, CA, U.S.A.). Also, droplet digital PCR (ddPCR) assay in the QX200 Droplet Digital PCR system (Bio-Rad, Carlsbad, CA, U.S.A.) was employed to measure methylation extent at 13 selected CpG sites, which had been reported to show robust evidence for association with lipid traits in previous EWASs (Supplementary Methods, Table S3).^10^

Because methylation profiles reflect environmental effects, MRSs were created for lipid traits using weighted sums of beta values from 13 CpG sites.^24^ Here, the effect sizes from the previous multi-ancestry EWAS for lipid traits^10^ were utilized as external weights for each CpG probe.

### Statistical analysis

Pearson’s correlation coefficient, *r*, was calculated to measure the strength and direction of the linear relationship between actual lipid levels and those predicted by the individual risk score models. To develop a prediction model, the relationship between predictor variables (i.e., PRS, MRS, and/or PRS+MRS) and outcome variables (i.e., plasma lipid levels) was examined in this study by two analytical methods: quantile binning and multivariable regression analysis. Quantile binning was used to convert an individual’s PRS to the absolute scale (i.e., plasma lipid value) as previously reported.^25^ In situations where the correctness of the linearity assumption is undetermined, quantile binning is essential to correct LDL-C changes due to rare functional variants and to combine them with the MRS at the individual level in a representative sample. Multivariable regression analyses, on the other hand, were performed to quantify the predictive utility of predictors (i.e., risk scores) at the group level separately by (sub)group, and the coefficient of determination (denoted *R*^2^) was used to provide a measure of how well the observed outcomes are approximated by the regression predictors in the tested population. The imputed value for lipid-lowering therapy was used in analyses including PRS when analyzing LDL-C.

No formal sample size calculations were performed since both BIO-CVD and NCGM Biobank are ongoing studies and there is no generally accepted approach to estimating the sample size needed to develop a prediction model like this study.

Significant results are shown as mean ± SE with *P* <0.05 unless otherwise stated.

## Results

### Imputation of baseline LDL-C level

This study recruited adult patients with high LDL-C and CAD at NCGM; of 315 CAD patients, 157 had a clear history of myocardial infarction, but 158 did not (Table 1). A considerable part of the participants had been prescribed statins, with 19% in the high LDL-C subgroup and 70% in the CAD subgroup (Table 1). For statin, irrespective of its type and dose, the average percent reduction in LDL-C from baseline (or pre-treatment) was approximately 40%. Also, for ezetimibe, the percent reduction in LDL-C was 22% (observation in a single patient) (Fig. S1). Hence, we used these values (40% for statins and 20% for ezetimibe) to impute pre-treatment LDL-C measurements for patients on lipid-lowering therapy.

### Rare variants in five FH-related genes

We identified 1,762 variants in five FH-related genes, of which 58 were rare (MAF <0.01) variants with functional significance. The variants (30 in *APOB*, 13 in *LDLR*, 7 in *PCSK9*, 1 in *LDLRAP1,* and 7 in *APOE*) could be classified into three categories—2 disruptive (frameshift and splice-donor each), 13 damaging, and 43 other non-synonymous variants (Table 2 and Table S2)—according to criteria described in the Methods. In a CAD patient, we also identified a gain-of-function type variant for *PCSK9* (indel, TGCCAGCGCCT/-), which appeared to be protective against CAD by exerting LDL-C lowering effects (Table 2). None of the identified rare variants overlapped with variants reported to affect response to statin therapy.^26^

**Table 2.** A list of functional variants identified by target resequencing of FH-related genes.

| Chr:Pos<br>(GRCh37,<br>hg19) | SNP | Ref/Alt allele | Gene<br>Name | Sequence<br>ontology <sup>a</sup> | Clinical<br>Signifi-<br>cance <sup>b</sup> | N of 6,<br>Predicted<br>as<br>Deleterious<br><sub>c</sub> | FATHM<br>M MKL<br>Coding<br>Pred (C) <sup>d</sup> | LOVD<br>classifi-<br>cation <sup>e</sup> | Alt allele_freq. | | $\chi^2$ test,<br>P-value | gnomAD<br>PopMax <sup>f</sup> | LDL-C<br>(mg/dl) | Notes |
| --- | --- | --- | --- | --- | --- | --- | --- | --- | --- | --- | --- | --- | --- | --- |
|  |  |  |  |  |  |  |  |  | Control<br>(ToMMo;<br>N = 8380) | HL+CAD<br>(N = 610) |  |  |  |  |
| Disruptive variants |  |  |  |  |  |  |  |  |  |  |  |  |  |  |
| 19:11215971 | rs879254510 | -/C | LDLR | frameshift | P/LP | — | — | P (ACGS) | N/A | 0.0008 | 2E-04 | N/A | 290 | D131fs |
| 19:11227676 | rs778408161 | T/C | LDLR | splice_<br>donor | P/LP | 2 of 6 | Damaging | LP<br>(ACGS) | 6E-05 | 0.0008 | 0.015 | 7E-06 | 271 | Disruptive (FH_Niigata) |
| Damaging variants |  |  |  |  |  |  |  |  |  |  |  |  |  |  |
| 19:11217342 | rs875989907 | G/A | LDLR | missense | P/LP | 6 of 6 | Damaging | LP | N/A | 0.0008 | 2E-04 | 6E-05 | 233 | GnomAD_exome (Asian);<br>3/49010 |
| 19:11221399 | rs879254753 | T/A | LDLR | missense | P/LP | 6 of 6 | Damaging | LP | 6E-05 | 0.0008 | 0.015 | N/A | 248 | Damaging (FH_Wakayama) |
| 19:11224019 | N/A | G/A | LDLR | missense | LP | 6 of 6 | Damaging | LP | 6E-05 | 0.0008 | 0.015 | N/A | 225 |  |
| 19:11226885 | rs746959386 | C/G | LDLR | missense | P/LP | 5 of 6 | Damaging | LP | 0.0003 | 0.0016 | 0.022 | 1E-04 | 189<br>± 57 |  |
| 19:11213368 | N/A | C/A | LDLR | missense | N/A | 6 of 6 | Damaging | N/A | N/A | 0.0008 | 2E-04 | N/A | 210 | newly identified |
| 19:11213417 | rs749038326 | G/A | LDLR | missense | Conflict-<br>ing<br>(LP/VUS<br>/LB*) | 6 of 6 | Damaging | P, LP | N/A | 0.0008 | 2E-04 | 3E-04 | 196 | GnomAD_exome (Asian);<br>14/49010 |
| 19:11216066 | N/A | C/T | LDLR | missense | N/A | 6 of 6 | Damaging | P (ACGS) | N/A | 0.0008 | 2E-04 | N/A | 212 | c.466_484dup |
| 19:11227613 | rs201102492 | G/A | LDLR | missense | Conflict-<br>ing<br>(P/LP/V<br>US) | 6 of 6 | Damaging | LP<br>(ACGS) | 6E-05 | 0.0008 | 0.015 | 2E-04 | 165 |  |
| 2:21228367 | N/A | G/T | APOB | missense | N/A | 4 of 6 | Damaging | N/A | N/A | 0.0008 | 2E-04 | N/A | 202 | newly identified |
| 2:21233912 | rs759277505 | G/A | APOB | missense | N/A | 5 of 6 | Damaging | N/A | 0.00018 | 0.0016 | 0.0004 | 1E-05 | 187<br>± 17 | A subject possessing<br>rs752849346 (LDLRAP1) and<br>rs564427867 (PCSK9)<br>concomitantly. |
| 2:21236137 | rs780170292 | C/T | APOB | missense | VUS | 5 of 6 | Damaging | N/A | 6E-05 | 0.0008 | 0.015 | 3E-04 | 170 |  |
| 2:21225680 | rs375471570 | G/A | APOB | missense | VUS | 4 of 6 | Damaging | N/A | 6E-05 | 0.0008 | 0.015 | 1E-04 | 210 |  |
| 2:21229473 | rs199689957 | C/T | APOB | missense | VUS | 2 of 6 | Damaging | N/A | N/A | 0.0008 | 2E-04 | 2E-04 | 293 | GnomAD_exome (Asian);<br>0/49006.<br>A subject possessing<br>rs564427867 (PCSK9)<br>concomitantly. |
| **Gain-of-Function variants** |  |  |  |  |  |  |  |  |  |  |  |  |  |  |
| 1:55521767 | N/A | TGCCAGCGCCT/- | PCSK9 | frameshift |  | — | — | — | N/A | 0.0008 | 2E-04 | N/A | 46 | NP_777596.2:p.Cys301Glyfs<br>Ter?<br>newly identified |

<sup>a</sup>The highest priority ontology found among the variant–transcript interactions. The terms used are the standard feature descriptions given by the Sequence Ontology Project.
<sup>b</sup>Clinical significance is the consensus interpretation of the submissions for all conditions for this variant. Primarily based on ACMG Classifications: P, pathogenic; LP, likely pathogenic; VUS, variant of uncertain significance; LB, likely benign; B, benign.
<sup>c</sup>Variant effect predictor tools tested are: SIFT, PolyPhen2, Mutation Taster, Mutation Assessor, FATHMM, and FATHMM-MKL.
<sup>d</sup>If a FATHMM-MKL coding score (PMID: 25583119) is >0.5 (or rank score >0.28317), the corresponding nsSNV is predicted as "DAMAGING"; otherwise it is predicted as "TOLERATED".
<sup>e</sup>Variant classification and interpretation by the Association for Clinical Genomic Science (ACGS) is shown when applicable. For abbreviations, refer to the ACMG Classifications above.
<sup>f</sup>PopMax refers to the gnomAD subpopulation with the highest allele frequency
\*Despite one submission (SCV000503122.1) indicating LB, the others claim either P (6 submissions) or LP (3 submissions) for the pathogenicity of this variant.

We evaluated the effect sizes of rare FH-related gene variants compared to a reference group of non-carriers by category to calculate the net increase in LDL-C caused by individual rare variants (see Methods). LDL-C levels increased significantly (P<0.05) in all three categories compared to the reference group: plus 106, 42, and 13 mg/dl for disruptive, damaging, and non-synonymous rare variants, respectively (Fig. 2A). The order of increase was consistent with the predicted pathogenicity of identified rare variants (Fig. 2B) and also with results from another study in Japanese.^22^

**Fig. 2.**
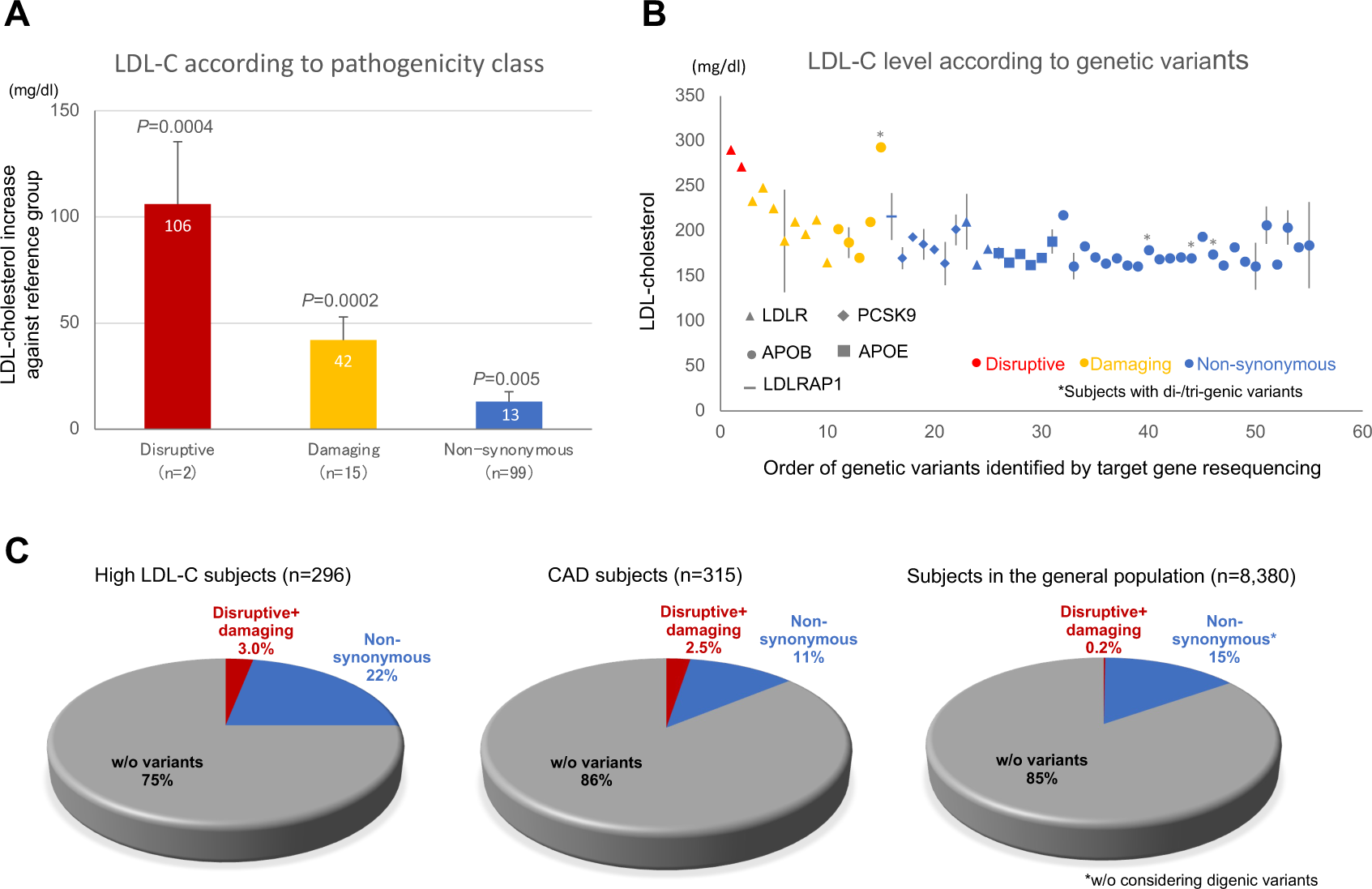
Impacts of rare FH-related gene variants on LDL-C level and their prevalence. An LDL-C increase against the reference group (BIO-CVD subjects without notable rare variants, *N* = 305) is estimated for three categories, i.e., disruptive, damaging, and non-synonymous (see Methods about the classification), of 5 FH-related gene variants (**A**), based on LDL-C level according to rare variants that were identified by target gene resequencing (**B**). **C:** The percentage of ‘disrup+ve + damaging’ variant carriers is shown for popula+ons with different morbidity statuses: high LDL-C (leb) and CAD (middle) subgroups and a general popula+on (right; TMM cohort) of Japanese descent.

The ratio of carriers for ‘disruptive + damaging’ variants was significantly (*P* <1×10^−15^) higher among CAD and high LDL-C subjects (ratio=0.025 and 0.03, respectively) than in the general population, TMM (ratio=0.002). (Fig. 2C).

### PRS and composite genetic risk prediction for LDL-C

By use of the standardized PRS_LDL-C_ decile classes, we found that PRS_LDL-C_ was shifted towards higher (from 7^th^ to 10^th^) decile classes among high LDL-C subjects, whereas PRS_LDL-C_ distribution was protruded in the 9^th^ decile class among CAD subjects (Fig. 3C).

**Fig. 3.**
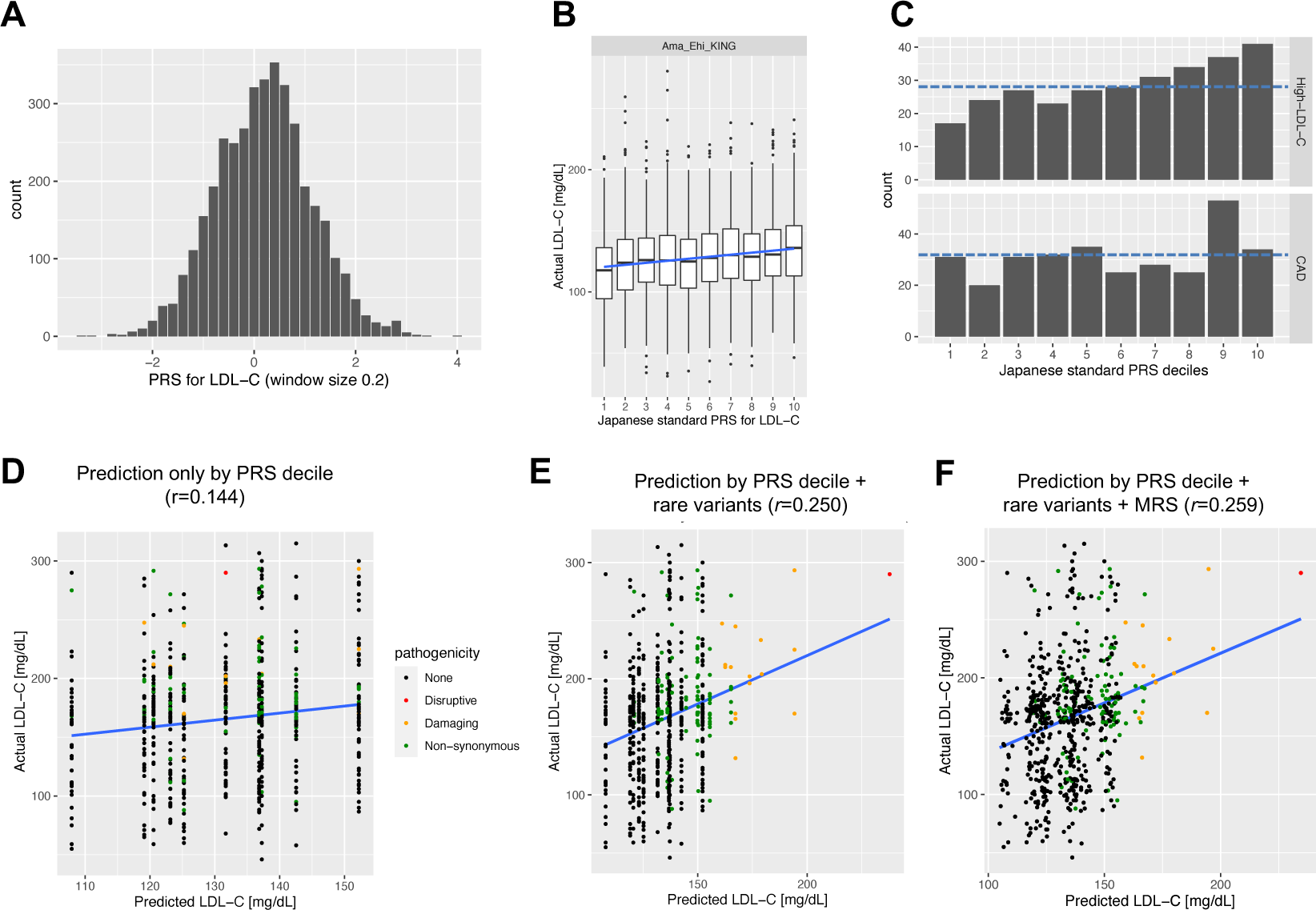
Development of a conversion formula from PRS to LDL-C value for PRS-based risk predicJon. First, to define the standardized PRS_LDL-C_ decile classes, data for three population-based cohorts (Kita-Nagoya, Amagasaki, and Ehime) were combined; distribution of PRS_LDL-C_ (**A**) and actual LDL-C for each PRS_LDL-C_ decile class thus standardized (**B**). Then, data for another hospital-based study cohort (NCGM hospital cohort) were used to convert PRS to LDL-C value for the individuals newly recruited at NCGM for rare variant search besides PRS and MRS calculations (see also Fig. S2). **C:** PRS_LDL-C_ distribu+on is stra+fied based on standardized PRS_LDL-C_ decile classes for high LDL-C (top) and CAD (boiom) subgroups at NCGM. Correlations are shown between actual LDL-C levels and those predicted by the standardized PRS_LDL-C_ decile class alone (**D**), composite genetic risk (**E**), and composite genetic risk + MRS (**F**) in newly recruited individuals at NCGM (BIO-CVD + NCGM Biobank) with a complete dataset.

We examined the correlation between predicted and actual LDL-C levels in newly recruited individuals at NCGM, based on standardized PRS_LDL-C_ decile and estimates of LDL-C increase by rare variant category in Japanese. (Fig. 3D,E). The correlation was stronger for composite genetic risk prediction with both PRS_LDL-C_ and rare variant effects (middle plot; *r*=0.261, *P* = 1.7×10^−11^) than conventional genetic risk prediction with PRS_LDL-C_ alone (left plot; *r*=0.151, *P* = 1.2×10^−4^).

### Impacts of PRSs and MRSs on lipid traits

Each subgroup (Table 1) had unimodal distributions of HDL-C, triglycerides, and BMI, distinct from LDL-C (Fig. S4). Between high LDL-C and CAD subgroups, there were significant differences in BMI (*P*=0.004) and HDL-C (*P*=9.2×10^−18^) in addition to LDL-C (*P*=5.8×10^−158^).

We confirmed the consistency (*R*^2^=0.34—0.86) of two DNA methylation analytical methods (EPIC array and ddPCR; see Fig. S5). Then, we used ddPCR to measure the methylation levels of 13 CpGs (see Table S3) and found that the effect sizes of the CpG-trait association were well correlated with previous multi-ethnic EWAS (Fig. 4A). Furthermore, there were significant differences (*P*=0.0045 for LDL, *P*=3.5×10^−7^ for HDL) in the MRS distribution between the high LDL-C and CAD subgroups (Fig. S6), consistent with phenotypic differences between the subgroups.

**Fig. 4.**
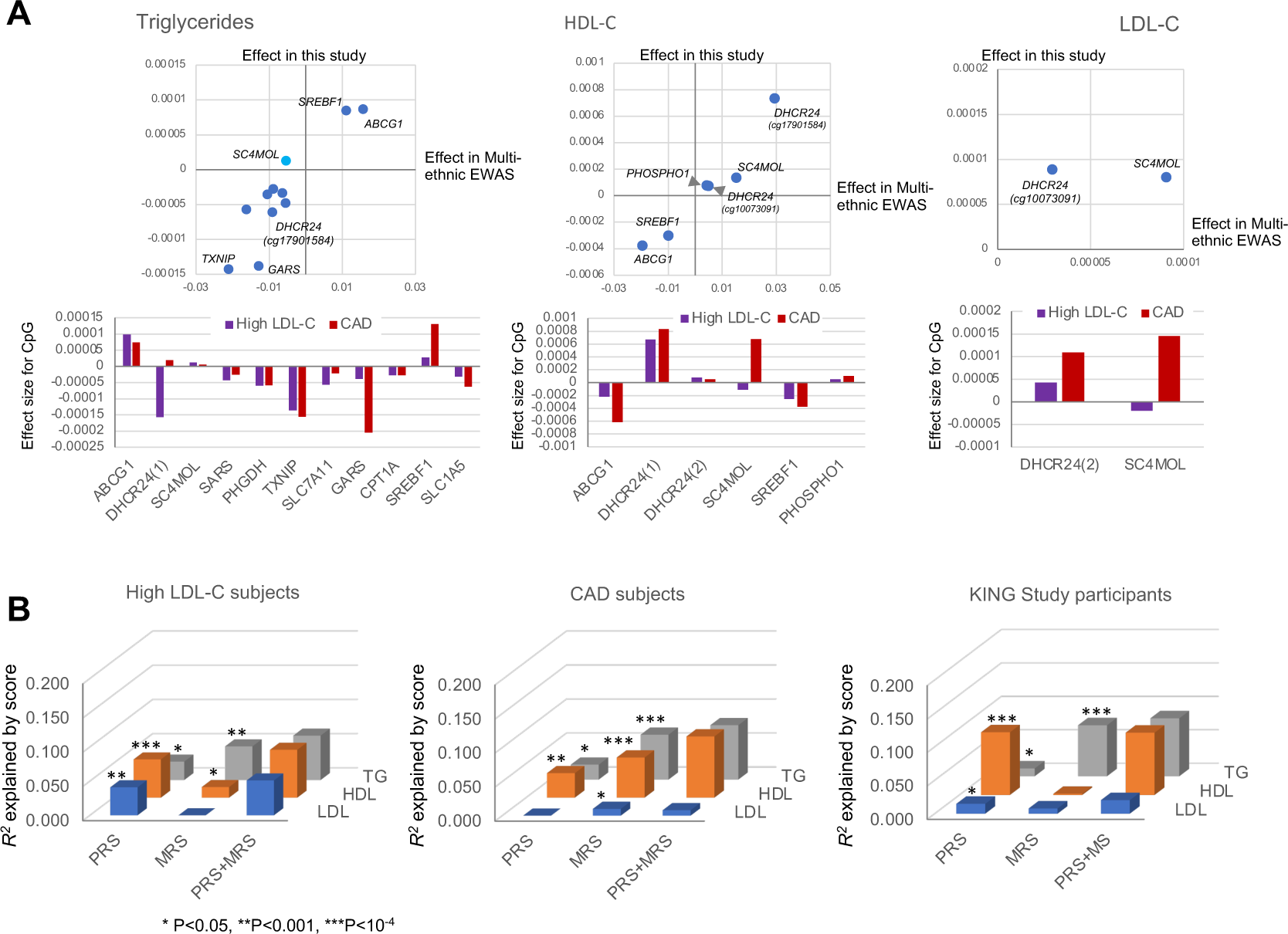
CpG-trait associaJon and predicJon of lipid traits by risk scores. **A:** Effect sizes for CpG-trait associa+on are shown for triglycerides (leb; at 11 CpGs), HDL-C (middle; at 6 CpGs), and LDL-C (right; at 2 CpGs), where comparisons are made between this study and previous mul+-ethnic EWAS^10^ (top panels), and between high LDL-C and CAD subgroups (boiom panels). **B:** Impacts of two risk scores, PRS and MRS, on lipid traits are es+mated by *R*^2^ in high LDL-C (leb) and CAD (middle) subgroups and a general popula+on (right; KING study cohort) of Japanese descent separately. \**P*<0.05, \*\**P*<0.001, \*\*\**P*<10^−4^.

When the CpGs were analyzed individually, the CpG-trait association was more robust in the CAD subgroup compared to the high LDL-C subgroup for lipid traits (Fig. 4A). This was supported by the results of the MRS-trait association, which involves the cumulative effects of top-hit CpGs previously reported for each lipid trait (Fig. 4B and Table S4). Contrarily, the impact of PRSs on lipid traits tended to be smaller in the CAD subgroup than in the high LDL-C subgroup. The two risk scores were also explanatory factors for the variance of lipid traits in the general population (as represented by the KING study cohort). At the same time, there were some differences in their relative influence compared to the high LDL-C and CAD subgroups. Overall, within each subgroup or population, the effects of PRSs and MRSs on lipid traits were additive (Fig. 4B). Although it remains a preliminary finding given the modest sample size in the current non-CAD subgroup, the strength of association between the risk scores and lipid traits appeared to be influenced by CAD status or its related factors (Table S4).

### Predictability of lipid traits by risk score models

We compared correlation strengths between predicted and measured lipid trait values using various risk score models to assess individual predictability (Fig.3D-F and Fig. S7). The model incorporating PRS and MRS (plus rare variants for LDL-C) showed a higher correlation than the one using only one risk score, supporting its efficacy in enhancing predictability.

## Discussion

This study, focusing on cardiovascular risk, evaluates the relative contribution of genetic predisposition and DNA methylation to plasma lipid levels, by developing a prediction model for dyslipidemia. Our analysis newly demonstrates two key points. First, concerning genetic predisposition, the effect sizes of rare FH-related gene variants are generally more prominent than the integrated effect of common variants (i.e., PRS). Combining the two can increase the accuracy of predicting an individual’s genetic predisposition for LDL-C. Second, DNA methylation, which is assumed to reflect an individual’s history of environmental exposure, interacts with genetic predisposition to affect an individual’s blood lipid levels. Therefore, combining DNA methylation and genetic predisposition can enhance the accuracy of predicting optimal treatment.

As a first step in this study, we have successfully developed an algorithm to predict composite genetic risk for high LDL-C levels. Prioritizing the detection of rare pathogenic variants in population screening, as well as the concurrent use of PRS is a topic of active discussion for genomic conditions that are actionable and adult-onset.^27^ Thus, a study has shown a risk prediction model that combines rare pathogenic variants, PRS, and individual risk factor variables for breast cancer (i.e., dichotomous trait).^28^ However, this has not been previously known for lipids, a quantitative trait. The heritability of blood lipid levels, based on SNPs, is estimated to be relatively high (23.3% for LDL-C, 30.5% for triglycerides, and 32.8% for HDL-C); however, the susceptibility loci identified so far can only partially explain it.^29^ Among the lipid traits, causative gene variants with a strong genetic influence, or monogenic mutations are known principally for LDL-C and are applied in current clinical practice for the genetic diagnosis of FH. In a population-based sample of European descent, only 2% of those with severe hypercholesterolemia (LDL-C > 183 mg/dl) were reported to carry a monogenic mutation, and 23% had a high PRS (top 5^th^ percentile) for LDL-C, indicating the importance of assessing both monogenic and polygenic models simultaneously.^30^ On the other hand, in a hospital-based setting, only about 40% of patients clinically diagnosed with FH were estimated to have an FH-causing rare variant, with the remainder likely to be polygenic in origin.^31^ Genetic effects on LDL-C levels have also been studied in relation to myocardial infarction susceptibility, with similar results to the present study for the FH-related genes *LDLR* and *PCSK9*.^22^ Accordingly, the newly developed algorithm for predicting LDL-C levels is expected to optimize preventive therapy for CAD patients.

In addition to genetic predisposition, epigenetics, specifically DNA methylation, has attracted attention in mediating gene-environment interactions in cardiometabolic phenotypes. This is because nutritional and lifestyle factors considerably influence epigenetics.^6–8^ Also, EWAS has shown through longitudinal analyses that smoking and alcohol consumption are associated with individual differences in DNA methylation at many CpG sites in the genome,^6,7^ and that such an association between traits and CpG methylation is likely to reflect to a significant degree the reverse causation of cardiometabolic phenotypes, including obesity.^32^ Furthermore, it has recently been noted that GWAS and EWAS capture different aspects of the biology of complex traits.^33^ Thus, interest has been growing in applying the PRS approaches to DNA methylation data, the so-called MRS. Still, methodological challenges must be resolved in its construction and use as a biomarker for environmental exposures.^34^

Given these circumstances, we have evaluated lipid MRSs containing a list of 13-CpGs in combination with PRS or without PRS through the development of prediction models for individual trait values. We initially selected the 13 CpGs because they showed a strong association with any of the lipid traits in the multi-ethnic EWAS,^10^ but these CpG-trait associations were reported to extend beyond lipids, considerably overlapping with other cardiometabolic phenotypes such as blood pressure, alcohol intake, and liver fat (Table S5). This leads to a hypothesis that some upstream environmental determinant (e.g., diet or exercise) is responsible for the overlap. For lipids, the effect sizes for single CpG-trait association between the present study in Japanese and previous multi-ethnic EWAS^10^ correlated well (Fig. 4A), which corroborates the reported high correlation between Europeans and non-Europeans (e.g., African Americans).^10^ Although there are some differences between subgroups in the interindividual variance explained by the MRS, the MRS predicts lipid traits in an additive manner independent of the PRS, particularly prominent among CAD subjects (Figure 4B and Table S4). This is in accord with previous findings on BMI, suggesting that the MRS represents environmental effects.^24^

While the clinical benefits of LDL-C lowering are widely acknowledged for the prevention of atherosclerotic cardiovascular diseases, a residual risk beyond LDL-C, including higher plasma levels of triglycerides and triglyceride-related apolipoproteins, has become recognized. In general, management of hypertriglyceridemia starts with lifestyle modification, and the use of drugs may be considered in high-risk individuals with limited benefits from lifestyle modification. This necessitates the encouragement of patient adherence to lifestyle changes or drug regimens.^1^

We envision that the combination of PRS (or, more preferably, composite genetic risk prediction) and MRS will contribute to a tailored approach to the management of dyslipidemia and even CAD, as emphasized by the clinical practice guidelines.^1,4^ In view of the considerable individual variability in not only dietary habits but also the LDL-C response to dietary and drug treatment,^35^ the use of our risk prediction index (exemplified in Fig. S8) in clinical practice may aid patient-doctor communication, facilitating adherence to and optimization of lipid treatment.

There are strengths and limitations in this study. First, we investigated the impact of genetic and DNA methylation variations on dyslipidemia in two risk groups (high LDL-C and CAD subjects), in contrast to the general population and non-CAD subjects. It is a strength that this information can be used to optimize treatment for primary (high LDL-C) and secondary (CAD) prevention of atherosclerotic cardiovascular disease by considering the relative contributions of genetics and environmental exposure. However, given age differences, individual differences in various lifestyle habits, and the impact of drug therapy on blood lipid measurements, data collected over time (including pretreatment measurements) in larger populations should be used to improve the accuracy of dyslipidemia risk prediction. Second, although we used 13 CpGs selected from top-hit CpGs in the multi-ethnic EWAS of lipids,^10^ the method for determining a set of CpGs for the MRS is still largely undefined. To improve the precision and practicality of the prediction model, an increased number of methylation sites for MRS and standard procedures for optimal weighting and clamping of target CpG sites are needed.^14^ Third, although the interaction term was included in multivariate regression analysis, potential interactions between MRS and PRS remain to be deeply explored. Fourth, combining rare FH-related gene variants with PRS can improve the predictability of LDL-C. Still, it must be more feasible and cost-effective to achieve similar predictability for HDL-C and triglycerides in clinical practice. Nevertheless, as the cost of sequencing decreases and genetic variation annotation becomes more manageable, a similar approach for the latter two lipid traits will be justifiable. Fifth, we utilized Japanese original reference panels with individual-level data for SNPs and the phenotypic trait of interest to standardize the PRS, which requires an ancestry-matched reference.^13^ For MRS, external weights from the multi-ethnic EWAS^10^ were used to calculate it, after confirming a high correlation between ethnic groups. However, further investigation is necessary to improve the generalizability of our risk prediction index. Sixth, given the limited number of CAD patients in the general Japanese population, the current findings need to be further validated and replicated with a larger, more representative population independently.

In summary, we conducted an integrative assessment towards precision medicine for dyslipidemia. Our data provided proof-of-concept that an individual’s dyslipidemia treatment regimen could be determined more precisely by the relative contribution of genetic predisposition and DNA methylation levels, which reflect past environmental exposures to a significant degree.

## Acknowledgments

We thank the Research Institute, NCGM staff for their technical assistance with DNA preparation and epigenetic analysis.

## Data availability

All data produced in the present study are available upon reasonable request to the authors.

