## Supplemental material for "Simultaneous assessment of genetic and epigenetic contributions to plasma lipid levels with respect to cardiovascular risk"

Fumihiko Takeuchi *et al*.

**SUPPLEMENTARY METHODS**

### ***1. Study population***

In NCGM, adult patients of Japanese descent were recruited via two separate projects: a bioresource collection project focused on cardiovascular disease, named BIO-CVD, and the NCGM Biobank project. They were then categorized into three subgroups based on their LDL-C levels and coronary artery disease (CAD) status: high LDL-C (≥160 mg/dl) without CAD, CAD, and non-CAD. We made a detailed assessment of CAD for samples in BIO-CVD, of which 353 (306 CAD and 47 non-CAD) subjects were used for resequencing of 5 target genes related to familial hypercholesterolemia (FH) and GWAS array genotyping. Here, CAD was defined as previously reported.[1] Briefly, the CAD criteria included (i) a validated history of either myocardial infarction (MI) or coronary revascularization (coronary artery bypass grafting or percutaneous coronary intervention), or (ii) subjective symptoms of angina pectoris with 1 or more major coronary vessels showing ≥75% stenosis documented by coronary angiography. Also, we examined the NCGM Biobank samples showing an elevated level of plasma LDL cholesterol (LDL-C ≥160 mg/dl) (*N* = 300), which were chosen from patients visiting NCGM for treatment of chronic illness. Besides the genetic analyses, we used these BIO-CVD and NCGM Biobank samples for DNA methylation analysis.

Independent of participants in the two projects above, we used many individuals derived from the general or non-targeted population of Japanese descent for genetic and DNA methylation analyses as follows. For low-frequency (or rare) genetic variants, we utilized whole genome sequencing data of participants (*N* = 8.3K) in the Tohoku Medical Megabank (TMM) project, which is publicly available. For common single nucleotide variants (SNVs), we also utilized GWAS array genotyping data for participants in three population-based cohort studies—Kita-Nagoya (*N* = 2,437), Amagasaki (*N* = 550) and Ehime (*N* = 463)—plus a hospital-based cohort study at NCGM (named NCGM hospital cohort; *N* = 351) previously reported,[2,3] which we referred to hereafter as reference individuals (*N* = 3,801) in total. Furthermore, for DNA methylation signature, we utilized EWAS-array assayed data for part (*N* = 314) of the Kita-Nagoya Genomic Epidemiology (KING) Study samples previously reported.[4]

### ***2. Gene sequencing***

Mutations were screened in five FH-related genes (*LDLR*, *APOB*, *PCSK9*, *LDLRAP1,* and *APOE*) by next-generation sequencing with the KAPA HyperChoice Kit (KAPA BioSystems, Wilmington, MA, U.S.A.) on an Illumina MiSeq or NextSeq 500. This covered the targets with a median read depth of 50× for coding DNA sequences, exon-intron junctions, and untranslated regions of the target genes. Read depth exceeded 20× for >90% of the target overall. The sequences were aligned to a human genome reference sequence (GRCh37/hg19). SNVs and insertion/deletion (Indel) were called and annotated using software GATK (https://gatk.broadinstitute.org/), McCortex (https://github.com/mcveanlab/mccortex), and VarSeq (https://www.goldenhelix.com/products/ VarSeq/).

#### 3. *Genotyping*

In this study, 644 samples were newly genotyped with the Infinium OmniExpress-24 BeadChip (Illumina, San Diego, CA, USA). Besides, GWAS array genotyping data for reference individuals were utilized. Data cleaning and analysis were performed using PLINK (version 1.07), and the imputation of genotypes to the 1000 Genomes Phase 3 set was carried out using Minimac3 software (version 2.0.1) as previously reported.[2]

#### 4. *Rare variant annotation*

We retrieved rare (MAF <0.01), putatively pathogenic variants, including SNVs that cause non-synonymous, nonsense, or splice-site substitutions and are predicted to be deleterious by ClinVar, LOVD or *in silico* prediction algorithms in dbNSFP (SIFT, Polyphen2, MutationTaster, MutationAssessor, FATHMM, and FATHMM-MKL).[5] For the rare variants of putative functional significance, we arbitrarily classified them into three distinct categories—(1) disruptive (frameshift or splice-donor), (2) damaging, and (3) all other non-synonymous variants—similar to the previous study[6] with some modifications. That is, for damaging variants, we defined them principally by missense variants annotated as either pathogenic/likely pathogenic in the reputable database or deleterious with ≥4 (out of 6) prediction algorithms in dbNSFP besides nominal significance level ($P$ <0.05) in the comparison of MAF (by chi-squared test) between case and reference groups.

#### 5. *Target CpG selection*

We performed a literature search on lipid-related EWASs and generated an initial list of target CpG methylation sites that show robust and reproducible association with any lipid traits; the lipid-related phenotypes include lipoprotein subfraction,[7,8] plasma lipids,[9-14] postprandial lipemia,[15] statin use,[16] and metabolic syndrome components.[17] Subsequently, we selected 13 CpGs to show a strong association with any lipid traits in the multi-ethnic EWAS, i.e., CpG markers with a significant correlation between Europeans and non-Europeans (e.g., African Americans).[18] Here, cg12556569 in the *APOA5* gene was excluded from the list since its probes for measuring DNA methylation show clustered distributions due to the underlying SNP (rs10750097, which is a relatively common variant in both Europeans and Asians) in the probe.[19]

#### *6. Methylation risk score for lipid traits*

Methylation risk scores (MRSs) were created for lipid traits using the weighted sums of beta values from individuals' methylation intensities at 13 CpG sites. The weights were determined as the effect sizes, represented as Z-scores, derived from a previous multi-ancestry EWAS for lipid traits [SuppData2, MODEL4.ALL in the research article by Jhun et al.[18]]. For each CpG site, we standardized the methylation beta values across the study population. The standardized values were then weighted and summed to derive the *raw* MRS for each individual. Following this, we performed a further standardization process to obtain the *standardized* MRS. To convert the *standardized* MRS to units of mg/dL, we utilized linear regression, aligning the *standardized* MRS with lipid measurements. The resulting scaling factors are 1.83 for LDL-C, 4.33 for HDL-C, and 19.0 for TG, respectively.

**Table S1. Participant characteristics of reference groups**

| | Population-based cohort | | | NCGM hospital cohort |
|---|---|---|---|---|
| | Kita-Nagoya | Amagasaki | Ehime | |
| No. of individuals (F/M) | 2437 (1272 /1165) | 550 (173 /377) | 463 (277/186) | 351 (133/218) |
| Age, yr | 63.9 ± 6.3 | 46.5 ± 12.5 | 64.4 ± 10.2 | 69.4 ± 8.4 |
| BMI. kg/m$^2$ | 22.8 ± 2.9 | 22.8 ± 2.9 | 23.2 ± 3.0 | 24.4 ± 3.4 |
| LDL-C, mg/dl | 131.1 ± 32.0 (F) | 122.0 ± 29.2 (F) | 124.3 ± 30.7 | 113.2 ± 30.4 |
| HDL-C, mg/dl | 62.6 ± 16.2 | 58.7 ± 15.8 | 65.2 ± 16.5 | 55.4 ± 14.8 |
| Triglycerides, mg/dl | 113.3 ± 57.2 | 113.4 ± 69.4 | 106.6 ± 57.3 | 140.5 ± 83.4 |
| Smoking habit<br>Current / Past / Never | 333/489/1602<br>(13 missing) | 213/60/275<br>(2 missing) | 33/119/311 | 125/106/113<br>(7 missing) |
| Complication | | | | |
| Hypertension, n (%) | 1149 (47%) | 106 (19%) | 174 (38%) | 287 (82%) |
| Diabetes, n (%) | 262 (11%) | 37 (7%) | 46 (10%) | 260 (74%) |
| Dyslipidemia*, n (%) | 1049 (43%) | 150 (27%) | 211 (46%) | 132 (38%) |
| Statin, n (%) | — | — | 89 (19%) | 134 (38%) |
| CAD, n (%) | 62 (3%) | 8 (1%) | 15 (3%) | 128 (36%) |

*In many cases, dyslipidemia is diagnosed as hyper-LDL-cholesterolemia. Hyper-LDL-cholesterolemia is defined when basal LDL-C ≥140 mg/dl (irrespective of treatment with lipid-lowering drugs, statin and/or ezetimibe) has been indicated by medical records. Detailed information about statin use was not recorded in the Kita-Nagoya and Amagasaki study cohorts.

**Table S2. A list of non-synonymous variants identified by target resequencing of FH-related genes**

| Chr:Pos (GRCh37, hg19) | SNP | Ref/Alt allele | Gene Name | Sequence ontology [a] | Clinical significance [b] | N of 6, Predicted as Damaging [c] | FATHMM MKL Coding Pred (C) [d] | LOVD_classification [e] | Alt allele_freq. | | $\chi^2$ test, P-value | gnomAD PopMax [f] | LDL-C (mg/dl) | Notes |
|---|---|---|---|---|---|---|---|---|---|---|---|---|---|---|
| | | | | | | | | | Control (ToMMo; n=8380) | HL+CAD (n=610) | | | | |
| 1:25880491 | rs752849346 | C/T | LDLRAP1 | missense | VUS | 4 of 6 | Damaging | N/A | 0.00125 | 0.0033 | 0.067 | 1E-04 | 216 ± 26 | PMID: 20124734 |
| 1:55505520 | rs186669805 | G/A | PCSK9 | missense | Conflicting (VUS/LB) | 0 of 6 | Tolerated | P | 0.01325 | 0.0098 | 0.31 | 7E-04 | 170 ± 12 | PCSK9 V4I reported in multiple Japanese FH-affected individuals (PMID: 26374825, 27206942) |
| 1:55505581 | N/A | G/A | PCSK9 | missense | N/A | 1 of 6 | Tolerated | N/A | N/A | 0.0008 | 0.00021 | N/A | 193 | p.Gly(G)24Asp(D) |
| 1:55505604 | rs564427867 | G/A | PCSK9 | missense | Conflicting (P/VUS/LP) | 1 of 6 | Damaging | P | 0.01104 | 0.0082 | 0.35 | 0.001 | 185 ± 17 | PCSK9 E32K reported in FH-affected individuals |
| 1:55518408 | rs749573024 | G/A | PCSK9 | missense | Conflicting (VUS/LB) | 3 of 6 | Damaging | N/A | N/A | 0.0008 | 0.00021 | 7E-05 | 179 | p.Arg(R)248His(H); increased proteolytic activity (PMID: 29259136) |
| 1:55518452 | rs200146448 | G/A | PCSK9 | missense | Conflicting (VUS/LB) | 3 of 6 | Damaging | N/A | 0.00698 | 0.0049 | 0.40 | 6E-04 | 164 ± 24 | |
| 1:55529108 | rs143291739 | G/A | PCSK9 | missense | VUS | 0 of 6 | Tolerated | N/A | 0.00352 | 0.0033 | 0.89 | 3E-04 | 201 ± 17 | |
| 19:11215926 | rs201102461 | G/A | LDLR | missense | VUS | 3 of 6 | Damaging | LP | 0.00442 | 0.0025 | 0.31 | 0.002 | 210 ± 31 | known as FH Fukuoka or p.R94H (PMID: 12837857) |
| 19:11215977 | rs751519676 | G/A | LDLR | missense | N/A | 1 of 6 | Tolerated | N/A | 0.0003 | 0.0008 | 0.34 | 1E-04 | 162 | |
| 19:11216033 | rs763233960 | G/A | LDLR | missense | Conflicting (P/LB) | 2 of 6 | Damaging | LB (ACGS) | 0.00012 | 0.0008 | 0.07 | 2E-05 | 180 | |
| 19:45409136 | rs373985746 | G/A | APOE | missense | N/A | 0 of 6 | Tolerated | VUS | 0.00221 | 0.0049 | 0.061 | 0.004 | 176 ± 7 | |
| 19:45409900 | N/A | G/C | APOE | missense | N/A | 4 of 6 | Tolerated | N/A | 0.00036 | 0.0008 | 0.43 | N/A | 165 | |
| 19:45411034 | rs121918392 | G/A | APOE | missense | P | 2 of 6 | Damaging | N/A | 0.00215 | 0.0025 | 0.82 | 3E-04 | 175 ± 1 | APOE5 allele, E21K. PMID: 7586659, 12069856 |
| 19:45411064 | rs201672011 | G/A | APOE | missense | P | 1 of 6 | Damaging | VUS | 0.00012 | 0.0008 | 0.07 | 0.003 | 162 | APOE5 allele, E31K. PMID: 7586659, 12069856 |
| 19:45412062 | N/A | C/T | APOE | missense | N/A | 1 of 6 | Tolerated | N/A | N/A | 0.0008 | 0.00021 | N/A | 170 | |
| 19:45412337 | rs140808909 | G/A | APOE | missense | P | 3 of 6 | Tolerated | N/A | 0.00817 | 0.0057 | 0.36 | 0.0026 | 188 ± 13 | APOE7, cooccurrence with rs190853081. PMID: 33928131 |
| 19:45412340 | rs190853081 | G/A | APOE | missense | P | 3 of 6 | Damaging | N/A | 0.00817 | 0.0057 | 0.36 | 0.0026 | | APOE7, cooccurrence with rs140808909. PMID: 33928131 |
| 2:21225472 | N/A | T/A | APOB | missense | N/A | 0 of 6 | Tolerated | N/A | 0.00066 | 0.0008 | 0.83 | N/A | 218 | |
| 2:21225713 | rs570782024 | A/G | APOB | missense | Conflicting (LB/VUS/B) | 1 of 6 | Tolerated | N/A | 0.00304 | 0.0049 | 0.26 | 0.0022 | 161 ± 15 | |
| 2:21227149 | N/A | T/C | APOB | missense | N/A | 2 of 6 | Damaging | N/A | N/A | 0.0008 | 0.00021 | N/A | 183 | |
| 2:21229422 | rs13306192 | A/C | APOB | missense | VUS | 5 of 6 | Damaging | N/A | 0.00227 | 0.0008 | 0.29 | 3E-04 | 171 | |
| 2:21229716 | N/A | A/T | APOB | missense | N/A | 5 of 6 | Damaging | N/A | 0.00054 | 0.0008 | 0.69 | N/A | 164 | |
| 2:21230147 | N/A | C/G | APOB | missense | N/A | 1 of 6 | Tolerated | N/A | N/A | 0.0008 | 0.00021 | N/A | 170 | |
| 2:21230891 | rs141591543 | A/G | APOB | missense | VUS | 2 of 6 | Damaging | N/A | 0.00066 | 0.0008 | 0.83 | 1E-04 | 162 | |
| 2:21231337 | rs374556458 | T/A | APOB | missense | N/A | 0 of 6 | Tolerated | N/A | N/A | 0.0008 | 0.00021 | 2E-05 | 161 | GnomAD_exome (Asian); 0/49006. |
| 2:21231348 | N/A | C/T | APOB | missense | N/A | 0 of 6 | Tolerated | N/A | N/A | 0.0008 | 0.00021 | N/A | 179 | |
| 2:21231629 | rs201874707 | G/A | APOB | missense | VUS | 4 of 6 | Damaging | VUS | 0.00095 | 0.0008 | 0.88 | 6E-05 | 169 | |
| 2:21231639 | rs774720452 | T/C | APOB | missense | N/A | 1 of 6 | Damaging | N/A | 6E-05 | 0.0008 | 0.015 | N/A | 170 | |
| 2:21232011 | rs200231583 | T/G | APOB | missense | B/LB | 0 of 6 | Tolerated | N/A | 0.0003 | 0.0008 | 0.34 | 1E-04 | 171 | cooccurrence with rs201152495 |
| 2:21232016 | rs201152495 | T/A | APOB | missense | LB | 2 of 6 | Damaging | N/A | 0.0003 | 0.0008 | 0.34 | 1E-04 | 171 | cooccurrence with rs200231583 |
| 2:21232517 | rs140027955 | G/A | APOB | missense | Conflicting (LB/VUS) | 4 of 6 | Damaging | N/A | 0.0003 | 0.0008 | 0.34 | 0.002 | 170 | |
| 2:21232775 | N/A | A/C | APOB | missense | N/A | 1 of 6 | Tolerated | N/A | 0.00089 | 0.0008 | 0.93 | N/A | 194 | |
| 2:21233351 | N/A | T/A | APOB | missense | N/A | 2 of 6 | Damaging | N/A | N/A | 0.0008 | 0.00021 | N/A | 174 | |
| 2:21233503 | rs747979961 | T/G | APOB | missense | N/A | 0 of 6 | Tolerated | N/A | 0.00042 | 0.0008 | 0.52 | 6E-05 | 162 | |
| 2:21234963 | N/A | T/C | APOB | missense | N/A | 0 of 6 | Tolerated | N/A | N/A | 0.0008 | 0.00021 | N/A | 182 | |
| 2:21235265 | N/A | A/C | APOB | missense | N/A | 2 of 6 | Damaging | N/A | N/A | 0.0008 | 0.00021 | N/A | 166 | |
| 2:21236085 | rs13306187 | C/T | APOB | missense | Conflicting (B/LB/VUS) | 0 of 6 | Tolerated | B | 0.00465 | 0.0008 | 0.05 | 0.001 | 161 ± 26 | Interpretation based on variant allele frequency (too common for FH) |
| 2:21236105 | rs754597311 | G/T | APOB | missense | Conflicting (VUS/LB) | 0 of 6 | Tolerated | N/A | 0.0031 | 0.0049 | 0.28 | 5E-04 | 207 ± 21 | Interpretation based on variant allele frequency (too common for FH) |
| 2:21241962 | rs1663539785 | G/C | APOB | missense | VUS | 5 of 6 | Damaging | N/A | 0.00125 | 0.0008 | 0.68 | 3E-04 | 163 | |
| 2:21247843 | rs183950016 | G/T | APOB | missense | VUS | 4 of 6 | Damaging | N/A | 0.00292 | 0.0016 | 0.42 | 3E-04 | 204 ± 19 | |
| 2:21259972 | N/A | C/T | APOB | missense | N/A | 3 of 6 | Damaging | N/A | N/A | 0.0008 | 0.00021 | N/A | 182 | |
| 2:21266769 | rs1197289618 | GCGCAG/- | APOB | inframe_ deletion | N/A | — | — | VUS | 0.00394 | 0.0025 | 0.42 | 6E-05 | 184 ± 48 | |

[a] The highest priority ontology found among the variant–transcript interactions. The terms used are the standard feature descriptions given by the Sequence Ontology Project.

[b] Clinical significance is the consensus interpretation of the submissions for all conditions for this variant. Primarily based on ACMG Classifications: P, pathogenic; LP, likely pathogenic; VUS, variant of uncertain significance; LB, likely benign; B, benign.

[c] Variant effect predictor tools tested are: SIFT, PolyPhen2, Mutation Taster, Mutation Assessor, FATHMM and FATHMM-MKL.

[d] If a FATHMM-MKL coding score (PMID: 25583119) is >0.5 (or rank score >0.28317), the corresponding nsSNV is predicted as "DAMAGING"; otherwise it is predicted as "TOLERATED".

[e] Variant classification and interpretation by the Association for Clinical Genomic Science (ACGS) is shown when applicable. For abbreviations, refer to the ACMG Classifications above.

[f] PopMax refers to the gnomAD subpopulation with the highest allele frequency

**Table S3A. BMI–CpG association in the current study**

| Gene | CpG probe | BMI-CpG association | | | | | | | | | | |
|---|---|---|---|---|---|---|---|---|---|---|---|---|
| | | Combined sample | | | High LDL-C sample | | | CAD sample | | | *P het* | Transethnic EWAS* |
| | | BETA | SE | *P* | BETA | SE | *P* | BETA | SE | P | high LDL-C vs CAD | Effect, Discovery/ Replication |
| ABCG1 | cg06500161 | 0.0015 | 0.0004 | 0.0003 | 0.0017 | 0.0006 | 0.004 | 0.0011 | 0.0006 | 0.05 | 0.45 | 34.8 / 19.9 |
| DHCR24 | cg17901584 | -0.002 | 0.001 | 0.14 | -0.002 | 0.0014 | 0.09 | -0.001 | 0.0015 | 0.41 | 0.55 | -13.7 / -7.3 |
| DHCR24 | cg10073091 | 0.0005 | 0.0003 | 0.08 | 0.0004 | 0.0004 | 0.39 | 0.0006 | 0.0005 | 0.17 | 0.71 | — |
| SC4MOL | cg05119988 | 0.0006 | 0.0006 | 0.34 | 0.001 | 0.0009 | 0.27 | 0.0003 | 0.0009 | 0.72 | 0.60 | — |
| SARS | cg03725309 | -9E-05 | 0.0003 | 0.78 | -3E-04 | 0.0005 | 0.53 | 2E-06 | 0.0004 | 0.997 | 0.63 | -24.8 / -8.2 |
| PHGDH | cg14476101 | -0.001 | 0.001 | 0.29 | 0.0002 | 0.0014 | 0.86 | -0.003 | 0.0014 | 0.07 | 0.15 | -14.5 / -8.9 |
| TXNIP | cg19693031 | -0.002 | 0.0007 | 0.004 | -0.003 | 0.001 | 0.003 | -8E-04 | 0.0009 | 0.39 | 0.11 | — |
| SLC7A11 | cg06690548 | -6E-04 | 0.0004 | 0.18 | -1E-04 | 0.0005 | 0.77 | -8E-04 | 0.0007 | 0.21 | 0.40 | -6.3 / -11.7 |
| GARS | cg21429551 | -9E-04 | 0.001 | 0.35 | -6E-04 | 0.0013 | 0.64 | -0.001 | 0.0015 | 0.43 | 0.77 | -6.9 / -4.7 |
| CPT1A | cg00574958 | -5E-04 | 0.0002 | 0.002 | -4E-04 | 0.0002 | 0.07 | -7E-04 | 0.0002 | 0.003 | 0.32 | -40.2 / -21.3 |
| SREBF1 | cg11024682 | 0.0023 | 0.0007 | 0.0004 | 0.0027 | 0.0009 | 0.002 | 0.0016 | 0.001 | 0.10 | 0.39 | 32.5 / 22.7 |
| PHOSPHO1 | cg02650017 | 7E-05 | 0.0001 | 0.57 | 0.0003 | 0.0002 | 0.10 | -1E-04 | 0.0002 | 0.45 | 0.09 | -28.6 / -15.2 |
| SLC1A5 | cg02711608 | -1E-04 | 0.0003 | 0.64 | 3E-05 | 0.0004 | 0.95 | -5E-04 | 0.0004 | 0.19 | 0.34 | -17.2 / -10.6 |

*Data are drawn from a study of transethnic EWAS for BMI (ref.24).

**Table S3B. Triglycerides–CpG association in the current study**

| Gene | CpG probe | Triglycerides-CpG association | | | | | | | | | | |
|---|---|---|---|---|---|---|---|---|---|---|---|---|
| | | Combined sample | | | High LDL-C sample | | | CAD sample | | | *P het* | Multi-ethnic EWAS* |
| | | BETA | SE | *P* | BETA | SE | *P* | BETA | SE | *P* | high LDL-C vs CAD | BETA |
| ABCG1 | cg06500161 | 9E-05 | 2E-05 | 2E-05 | 1E-04 | 3E-05 | 0.003 | 7E-05 | 3E-05 | 0.004 | 0.56 | 0.016 |
| DHCR24 | cg17901584 | -6E-05 | 5E-05 | 0.24 | -2E-04 | 8E-05 | 0.05 | 2E-05 | 7E-05 | 0.78 | 0.09 | -0.009 |
| DHCR24 | cg10073091 | 8E-06 | 2E-05 | 0.62 | -3E-05 | 2E-05 | 0.23 | 4E-05 | 2E-05 | 0.08 | 0.04 | — |
| SC4MOL | cg05119988 | 1E-05 | 3E-05 | 0.68 | 1E-05 | 5E-05 | 0.81 | 5E-06 | 4E-05 | 0.89 | 0.92 | -0.005 |
| SARS | cg03725309 | -3E-05 | 2E-05 | 0.03 | -4E-05 | 3E-05 | 0.10 | -3E-05 | 2E-05 | 0.15 | 0.60 | -0.006 |
| PHGDH | cg14476101 | -6E-05 | 5E-05 | 0.26 | -6E-05 | 8E-05 | 0.44 | -6E-05 | 7E-05 | 0.38 | 0.99 | -0.016 |
| TXNIP | cg19693031 | -1E-04 | 3E-05 | 2E-05 | -1E-04 | 5E-05 | 0.0098 | -2E-04 | 4E-05 | 0.0002 | 0.77 | -0.021 |
| SLC7A11 | cg06690548 | -4E-05 | 2E-05 | 0.08 | -6E-05 | 3E-05 | 0.04 | -2E-05 | 3E-05 | 0.49 | 0.38 | -0.011 |
| GARS | cg21429551 | -1E-04 | 5E-05 | 0.006 | -4E-05 | 7E-05 | 0.59 | -2E-04 | 7E-05 | 0.003 | 0.10 | -0.013 |
| CPT1A | cg00574958 | -3E-05 | 8E-06 | 0.0004 | -3E-05 | 1E-05 | 0.02 | -3E-05 | 1E-05 | 0.008 | 0.99 | -0.009 |
| SREBF1 | cg11024682 | 9E-05 | 3E-05 | 0.0098 | 3E-05 | 5E-05 | 0.58 | 0.0001 | 4E-05 | 0.004 | 0.12 | 0.011 |
| PHOSPHO1 | cg02650017 | 3E-06 | 6E-06 | 0.64 | -1E-06 | 9E-06 | 0.90 | 8E-06 | 8E-06 | 0.32 | 0.46 | — |
| SLC1A5 | cg02711608 | -5E-05 | 2E-05 | 0.002 | -3E-05 | 2E-05 | 0.20 | -6E-05 | 2E-05 | 0.0009 | 0.31 | -0.006 |

*Data are drawn from a study of multi-ethnic EWAS for lipids (ref.18).

**Table S3C. HDL-C–CpG association in the current study**

| Gene | CpG probe | HDL-C-CpG association | | | | | | | | | | |
|---|---|---|---|---|---|---|---|---|---|---|---|---|
| | | Combined sample | | | High LDL-C sample | | | CAD sample | | | $P_{het}$ | Multi-ethnic EWAS* |
| | | BETA | SE | *P* | BETA | SE | *P* | BETA | SE | P | high LDL-C vs CAD | BETA |
| ABCG1 | cg06500161 | -4E-04 | 1E-04 | 0.0002 | -2E-04 | 0.0001 | 0.09 | -6E-04 | 0.0002 | 0.0002 | 0.06 | -0.019 |
| DHCR24 | cg17901584 | 0.0007 | 0.0002 | 0.003 | 0.0007 | 0.0003 | 0.03 | 0.0008 | 0.0004 | 0.05 | 0.76 | 0.029 |
| DHCR24 | cg10073091 | 8E-05 | 8E-05 | 0.33 | 8E-05 | 9E-05 | 0.40 | 5E-05 | 0.0001 | 0.72 | 0.85 | 0.005 |
| SC4MOL | cg05119988 | 0.0001 | 0.0002 | 0.37 | -1E-04 | 0.0002 | 0.55 | 0.0007 | 0.0003 | 0.009 | 0.01 | 0.015 |
| SARS | cg03725309 | 5E-05 | 8E-05 | 0.53 | 8E-05 | 0.0001 | 0.46 | 1E-05 | 0.0001 | 0.90 | 0.69 | — |
| PHGDH | cg14476101 | -6E-05 | 0.0002 | 0.80 | 5E-05 | 0.0003 | 0.86 | -2E-04 | 0.0004 | 0.72 | 0.70 | — |
| TXNIP | cg19693031 | -1E-04 | 0.0002 | 0.37 | -1E-04 | 0.0002 | 0.60 | -2E-04 | 0.0003 | 0.55 | 0.88 | — |
| SLC7A11 | cg06690548 | 0.0001 | 0.0001 | 0.14 | 0.0002 | 0.0001 | 0.04 | 6E-05 | 0.0002 | 0.78 | 0.47 | — |
| GARS | cg21429551 | 0.0003 | 0.0002 | 0.19 | 0.0001 | 0.0003 | 0.62 | 0.0007 | 0.0004 | 0.14 | 0.33 | — |
| CPT1A | cg00574958 | 5E-06 | 4E-05 | 0.90 | 2E-05 | 4E-05 | 0.64 | -2E-05 | 7E-05 | 0.72 | 0.58 | — |
| SREBF1 | cg11024682 | -3E-04 | 0.0002 | 0.06 | -3E-04 | 0.0002 | 0.19 | -4E-04 | 0.0003 | 0.21 | 0.73 | -0.010 |
| PHOSPHO1 | cg02650017 | 8E-05 | 3E-05 | 0.007 | 5E-05 | 3E-05 | 0.13 | 0.0001 | 5E-05 | 0.04 | 0.41 | 0.004 |
| SLC1A5 | cg02711608 | -6E-06 | 7E-05 | 0.94 | -4E-06 | 1E-04 | 0.97 | 3E-05 | 0.0001 | 0.81 | 0.83 | — |

*Data are drawn from a study of multi-ethnic EWAS for lipids (ref.18).

**Table S3D. LDL-C–CpG association in the current study**

| Gene | CpG probe | LDL-C-CpG association | | | | | | | | | | |
|---|---|---|---|---|---|---|---|---|---|---|---|---|
| | | Combined sample | | | High LDL-C sample | | | CAD sample | | | $P_{het}$ | Multi-ethnic EWAS* |
| | | BETA | SE | P | BETA | SE | P | BETA | SE | P | high LDL-C vs CAD | BETA |
| ABCG1 | cg06500161 | -2E-05 | 6E-05 | 0.69 | 0.0002 | 0.0001 | 0.06 | -1E-04 | 7E-05 | 0.03 | 0.005 | — |
| DHCR24 | cg17901584 | 0.0003 | 0.0001 | 0.06 | 8E-05 | 0.0002 | 0.73 | 0.0003 | 0.0002 | 0.06 | 0.43 | — |
| DHCR24 | cg10073091 | 9E-05 | 4E-05 | 0.04 | 4E-05 | 7E-05 | 0.57 | 0.0001 | 5E-05 | 0.04 | 0.46 | 3E-05 |
| SC4MOL | cg05119988 | 8E-05 | 8E-05 | 0.34 | -2E-05 | 0.0001 | 0.90 | 0.0001 | 0.0001 | 0.15 | 0.36 | 9E-05 |
| SARS | cg03725309 | -2E-05 | 4E-05 | 0.57 | -4E-05 | 8E-05 | 0.59 | -3E-05 | 5E-05 | 0.55 | 0.86 | — |
| PHGDH | cg14476101 | 0.0001 | 0.0001 | 0.34 | 0.0002 | 0.0002 | 0.41 | 5E-05 | 0.0002 | 0.76 | 0.62 | — |
| TXNIP | cg19693031 | -1E-04 | 9E-05 | 0.28 | -3E-05 | 0.0002 | 0.86 | -1E-04 | 0.0001 | 0.26 | 0.63 | — |
| SLC7A11 | cg06690548 | 8E-05 | 6E-05 | 0.16 | 6E-05 | 8E-05 | 0.44 | 8E-05 | 8E-05 | 0.28 | 0.86 | — |
| GARS | cg21429551 | -5E-05 | 0.0001 | 0.71 | 0.0001 | 0.0002 | 0.57 | -2E-04 | 0.0002 | 0.38 | 0.32 | — |
| CPT1A | cg00574958 | -3E-06 | 2E-05 | 0.90 | -4E-05 | 4E-05 | 0.27 | 1E-05 | 3E-05 | 0.61 | 0.23 | — |
| SREBF1 | cg11024682 | 0.0002 | 9E-05 | 0.02 | 0.0003 | 0.0002 | 0.08 | 0.0001 | 0.0001 | 0.23 | 0.51 | — |
| PHOSPHO1 | cg02650017 | 2E-05 | 2E-05 | 0.21 | 9E-06 | 3E-05 | 0.75 | 3E-05 | 2E-05 | 0.15 | 0.57 | — |
| SLC1A5 | cg02711608 | -4E-05 | 4E-05 | 0.35 | -1E-04 | 8E-05 | 0.06 | -5E-06 | 5E-05 | 0.92 | 0.12 | — |

*Data are drawn from a study of multi-ethnic EWAS for lipids (ref.18).

**Table S4A. Association of PRS and MRS with triglycerides in a regression model**

| | PRS | | | MRS | | | PRS+MRS | Interaction [PRS×MRS] | |
|---|---|---|---|---|---|---|---|---|---|
| | Estimate (SE) | Pr(>\|t\|) | R-squared | Estimate (SE) | Pr(>\|t\|) | R-squared | R-squared | Estimate (SE) | Pr(>\|t\|) |
| High LDL-C | 6.932 (2.884) | 0.017 | 0.026 | 14.744 (4.162) | 4.6E-04 | 0.049 | 0.065 | 1.185 (3.220) | 0.713 |
| CAD | 7.903 (3.383) | 0.020 | 0.022 | 19.238 (4.340) | 1.3E-05 | 0.066 | 0.080 | 4.721 (3.165) | 0.137 |
| KING | 5.008 (2.355) | 0.034 | 0.012 | 15.826 (3.076) | 4.8E-07 | 0.076 | 0.087 | -2.158 (2.392) | 0.368 |
| non-CAD | -1.560 (5.274) | 0.768 | -0.005 | 7.082 (6.652) | 0.289 | 0.002 | -0.004 | -4.153 (5.541) | 0.455 |

We used a regression model to analyze the association between the independent variables (PRS, MRS, or PRS+MRS) and the dependent variable (triglycerides) addjusted for sex, age, smoking status and BMI. The independent variables were standardized to have a mean of 0 and a standard deviation of 1. To calculate the MRS for triglycerides, we multiplied the methylation extent measurements for 11 CpGs (listed in Table S3) with corresponding Z-scores from literature (Jhun et al., 10.1038/s41467-021-23899-y SuppData2, MODEL4.ALL) at each locus, and then integrated the products. Abbreviations used: PRS (polygenic risk score) and MRS (methylation risk score).

**Table S4B. Association of PRS and MRS with HDL-C in a regression model**

| | PRS | | | MRS | | | PRS+MRS | Interaction [PRS×MRS] | |
|---|---|---|---|---|---|---|---|---|---|
| | Estimate (SE) | Pr(>\|t\|) | R-squared | Estimate (SE) | Pr(>\|t\|) | R-squared | R-squared | Estimate (SE) | Pr(>\|t\|) |
| High LDL-C | 2.650 (0.652) | 6.4E-05 | 0.056 | 2.298 (1.033) | 0.027 | 0.016 | 0.070 | 0.302 (0.708) | 0.670 |
| CAD | 1.367 (0.411) | 9.9E-04 | 0.036 | 2.924 (0.678) | 2.2E-05 | 0.059 | 0.090 | 0.284 (0.440) | 0.519 |
| KING | 2.782 (0.490) | 3.1E-08 | 0.094 | 0.697 (0.788) | 0.377 | 0.002 | 0.093 | 0.378 (0.546) | 0.489 |
| non-CAD | 0.595 (0.678) | 0.381 | -0.006 | 3.908 (1.266) | 0.002 | 0.047 | 0.045 | 0.375 (0.733) | 0.610 |

We used a regression model to analyze the association between the independent variables (PRS, MRS, or PRS+MRS) and the dependent variable (HDL-C) addjusted for sex, age, smoking status and BMI. The independent variables were standardized to have a mean of 0 and a standard deviation of 1. To calculate the MRS for triglycerides, we multiplied the methylation extent measurements for 6 CpGs (listed in Table S3) with corresponding Z-scores from literature (Jhun et al., 10.1038/s41467-021-23899-y SuppData2, MODEL4.ALL) at each locus, and then integrated the products. Abbreviations used: PRS (polygenic risk score) and MRS (methylation risk score).

**Table S4C. Association of PRS and MRS with LDL-C in a regression model**

| | PRS | | | MRS | | | PRS+MRS | Interaction [PRS×MRS] | |
|---|---|---|---|---|---|---|---|---|---|
| Sub-group | Estimate (SE) | Pr(>\|t\|) | R-squared | Estimate (SE) | Pr(>\|t\|) | R-squared | R-squared | Estimate (SE) | Pr(>\|t\|) |
| High LDL-C | 8.577 (2.195) | 1.2E-04 | 0.041 | 0.134 (1.355) | 0.921 | -0.003 | 0.051 | -0.642 (2.197) | 0.770 |
| CAD | 1.639 (2.883) | 0.570 | -0.002 | 4.182 (1.744) | 0.017 | 0.01 | 0.008 | 3.470 (3.137) | 0.270 |
| KING | 3.906 (1.757) | 0.027 | 0.015 | 2.838 (1.712) | 0.098 | 0.008 | 0.02 | 0.023 (1.791) | 0.013 |
| non-CAD | 1.954 (3.978) | 0.624 | -0.005 | 2.039 (2.102) | 0.333 | 0.001 | -0.008 | 0.910 (3.656) | 0.804 |

We used a regression model to analyze the association between the independent variables (PRS, MRS, or PRS+MRS) and the dependent variable (LDL-C) addjusted for sex, age, smoking status and BMI. The independent variables were standardized to have a mean of 0 and a standard deviation of 1. LDL-C values with imputation were used for regression analysis except in the case of MRSs; LDL-C values without imputation were utilized for calculating MRSs in high LDL-C, CAD, and non-CAD subgroups to reflect the condition at the time of blood collection. For LDL-C in the KING Study, no imputation was performed. To calculate the MRS for LDL-C, we multiplied the methylation extent measurements for 2 CpGs (listed in Table S3) with corresponding Z-scores from literature (Jhun et al., 10.1038/s41467-021-23899-y SuppData2, MODEL4.ALL) at each locus, and then integrated the products.
Abbreviations: PRS, polygenic risk score; MRS, methylation risk score.

### Table S5. Overlap of DNA methylation-cardiometabolic trait association at 13 CpG sites tested in the present study

| | Trait | CRP | Blood pressure | Alcohol consumption | Hepatic fat | γ-glutamyl transferase | Gene expression | BMI/WC | Statin use | Triglycerides | HDL | LDL | T2D | Insulin | HbA1c | FPG |
|---|---|---|---|---|---|---|---|---|---|---|---|---|---|---|---|---|
| | PMID | 27955697 | 29198723 | 31789449 | 30936141 | 28624579 | 33752734 | 28002404/ 29099282 | 32033992 | 34183656 | 34183656 | 34183656 | 35169870 | 31197173 | 26643952 | 31197173 |
| Gene | No. of CpGs<br>CpG probe | 58 CpGs | 13 CpGs | 1414 CpGs | 22 CpGs | 4 CpGs | 148-1741 CpG-transcript pairs | 187 loci/ 82 CpGs | 5 CpGs | 216 CpGs | 110 CpGs | 39 CpGs | 64 CpGs | 18 CpGs | 1 CpG | 7 CpGs |
| ABCG1 | cg06500161 | | | | ○ | | ○ | ◎/null | ◎ | ◎ | ◎ | | ◎ | ◎ | | |
| DHCR24 | cg17901584 | | | | △ | | ◎ | ○/○ | ◎ | ◎ | ◎ | | | △ | | |
| DHCR24 | cg10073091 | | | | | | | | | | ○ | ○ | | | | |
| SC4MOL | cg05119988 | | | | | | | △/null | ○ | ○ | ○ | ○ | | | | |
| SARS | cg03725309 | △ | ○ | | △ | | | ○/null | | ◎ | | | | | | |
| PHGDH | cg14476101 | ○ | ◎ | ◎ | △ | △ | ○ | ◎/null | | ◎ | | | ○ | | | |
| TXNIP | cg19693031 | | ◎ | ◎ | ○ | | | null/○ | | ◎ | | | ◎ | | ○ | |
| SLC7A11 | cg06690548 | ○ | ◎ | ◎ | ○ | ◎ | | ◎/◎ | | ◎ | | | | | | ○ |
| GARS | cg21429551 | △ | ○ | ○ | △ | | | ○/null | | ◎ | | | | | | |
| CPT1A | cg00574958 | | ○ | ○ | ○ | | ◎ | ◎/◎ | | ◎ | | | ○ | ○ | | ○ |
| SREBF1 | cg11024682 | ○ | | | △ | △ | ○ | ◎/◎ | | ◎ | ○ | | ○ | △ | | △ |
| PHOSPHO1 | cg02650017 | ◎ | | | | | | ◎/null | | | ◎ | | ○ | | | |
| SLC1A5 | cg02711608 | ○ | ◎ | ◎ | △ | | ○ | ○/null | | ◎ | | | ○ | | | |

Positive hits (P<5E-8) are drawn from the EWAS Catalog (http://www.ewascatalog.org) as of 2023.0830.

Positive signals are arbitrarily categorized into three classes according to the P-value: triangle (△), 5E-8>P≥1E-10; circle (○), 1E-10>P≥1E-20; and double circle (◎), P<1E-20.

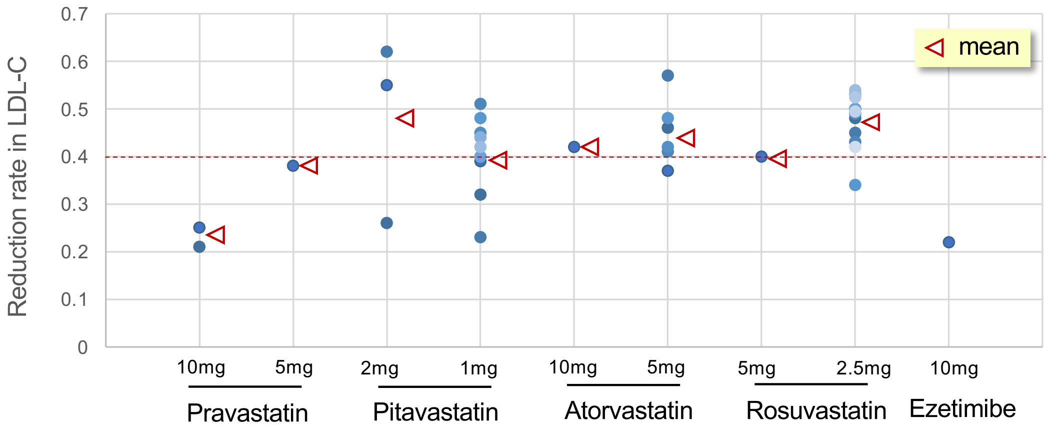

**Fig. S1: Reduction rate in LDL-C according to medication.**

The treatment-induced reduction rate in LDL-C was calculated in Japanese by a retrospective analysis of data from 36 individuals with high LDL-C in whom plasma LDL-C measurements were available both before and within 18 months after initiating statins and ezetimibe. The arrows indicate the mean of the reduction rate in each treatment group. In populations of European descent, while the treatment-induced reduction rate in LDL-C has been reported to be 0.2 to >0.5 according to dose and type of statins,[20] a reduction rate of 0.3 is generally used for imputation of LDL-C measured in those on statin treatment.[21,22] Given the difference between Asian and Western groups in the LDL-C lowering effect of statins (i.e., the dosages are significantly lower in Asian than in Western groups to attain a target effect),[23] we deemed 0.4 appropriate for statins in Japanese.

Fig.S2

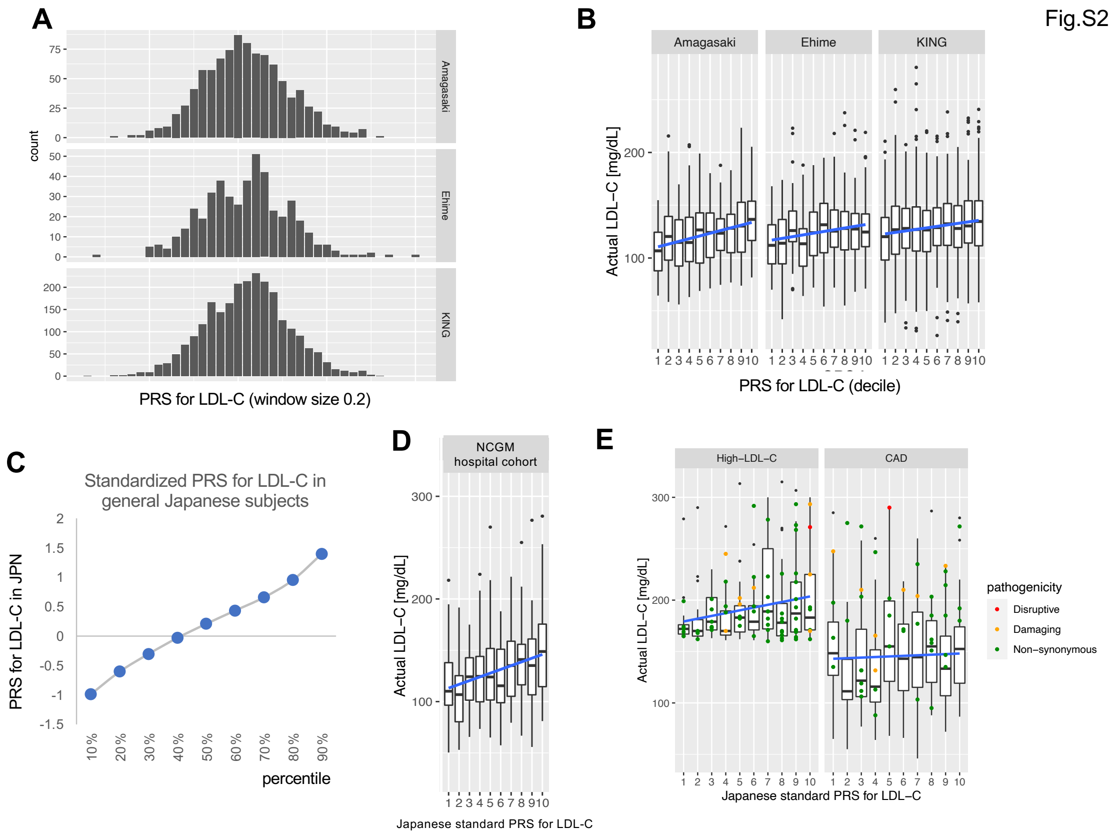

**Fig. S2: Estimating standardized impacts of a $PRS_{LDL\text{-}C}$ on plasma LDL-C level in Japanese.**

First, to define the standardized $PRS_{LDL\text{-}C}$ decile classes in Japanese (referred to as the 'Japanese standard PRS for LDL-C'), we utilized data from three population-based cohorts (Amagasaki, Ehime, and Kita-Nagoya [KING]), whose areas are somewhat geographically distant from each other in Japan. Next, to convert the $PRS_{LDL\text{-}C}$ from high LDL-C and CAD subgroups into LDL-C predictions, we used the mean of the LDL-C measurements from the NCGM hospital cohort (i.e., an external reference group) belonging to the same population as the above two subgroups recruited at NCGM. **A:** Histograms of $PRS_{LDL\text{-}C}$ in three cohorts. **B:** Box-and-whisker plots of LDL-C by $PRS_{LDL\text{-}C}$ decile in three cohorts. **C:** Line graphs of $PRS_{LDL\text{-}C}$ in Japanese at each decile point for the combined three cohort data. Box-and-whisker plots of LDL-C measurements from the NCGM hospital cohort (**D**) and the high LDL-C and CAD subgroups (**E**).

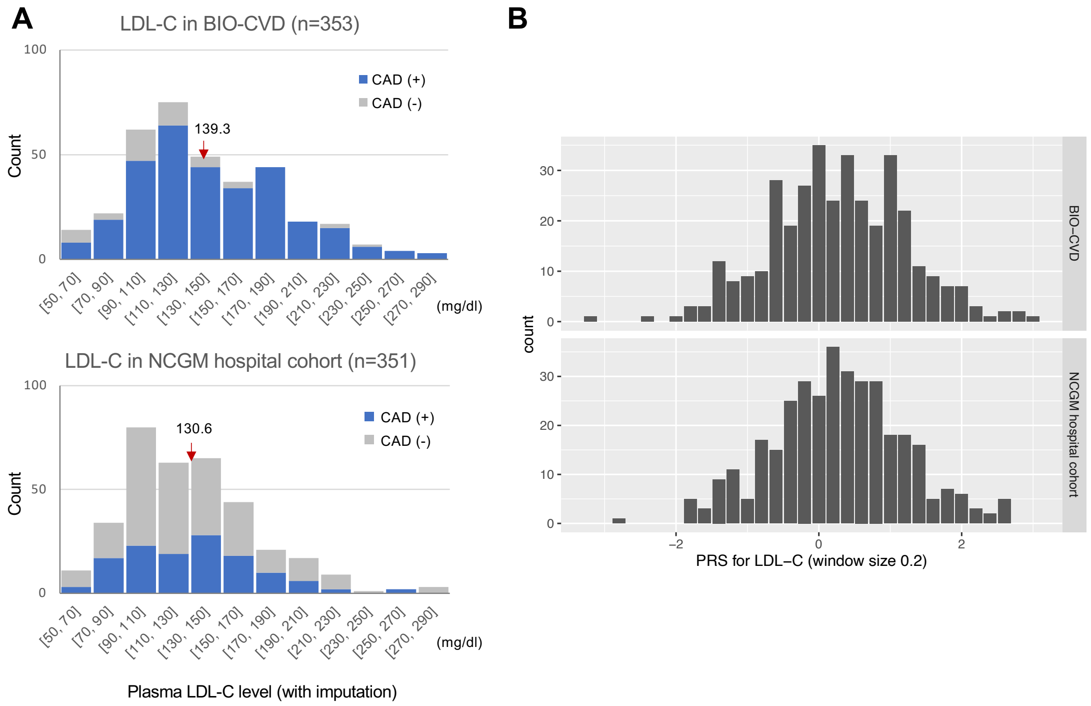

**Fig. S3: Distribution of LDL-C and PRS$_{LDL}$ in two hospital-based cohorts at NCGM.**

Histograms of plasma LDL-C level (**A**) and PRS$_{LDL\text{-}C}$ (**B**) in the BIO-CVD (top panels) and NCGM hospital cohort (bottom panels) projects. BIO-CVD is a bioresource collection project focused on cardiovascular disease, which includes 306 CAD and 47 non-CAD (see Fig. 1). On the other hand, the NCGM hospital cohort ($N$ = 351) was separately recruited without screening for cardiovascular disease. In the histograms of Fig. S3A**,** CAD (and non-CAD) subjects are colored in blue (and grey), and the arrows indicate the mean of LDL-C in individual project samples. Besides, non-CAD ($N$ = 125) subjects from BIO-CVD were added to GWAS array genotyping.

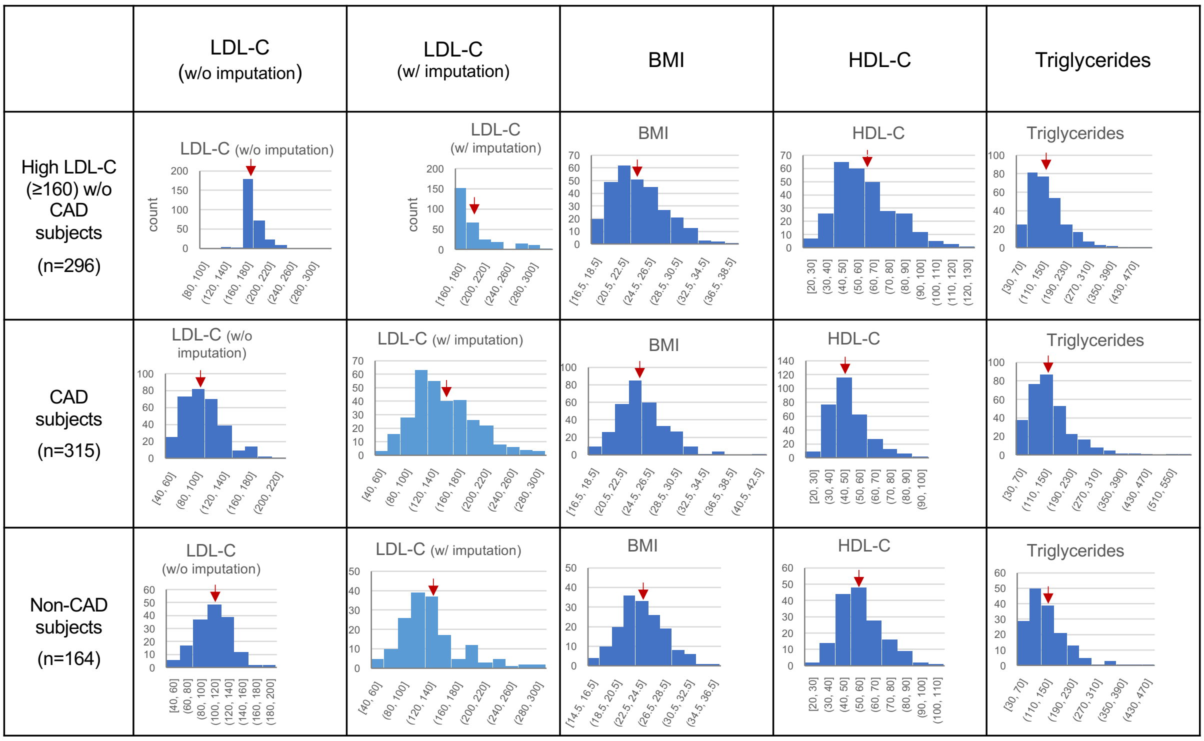

**Fig. S4: Distribution of metabolic traits in three subgroups newly recruited.**

Histograms of lipid traits and body mass index (BMI) are shown for three subgroups—high LDL-C (≥160 mg/dl), CAD (regardless of LDL-C level), and non-CAD individuals—newly recruited via two bioresource projects (BIO-CVD and NCGM Biobank). LDL-C was imputed for subjects taking lipid-lowering medications (statins or ezetimibe), and the imputed values (second column from left) were utilized in regression analysis except in the case of MRSs. The arrows indicate the mean of each metabolic trait in individual study groups.

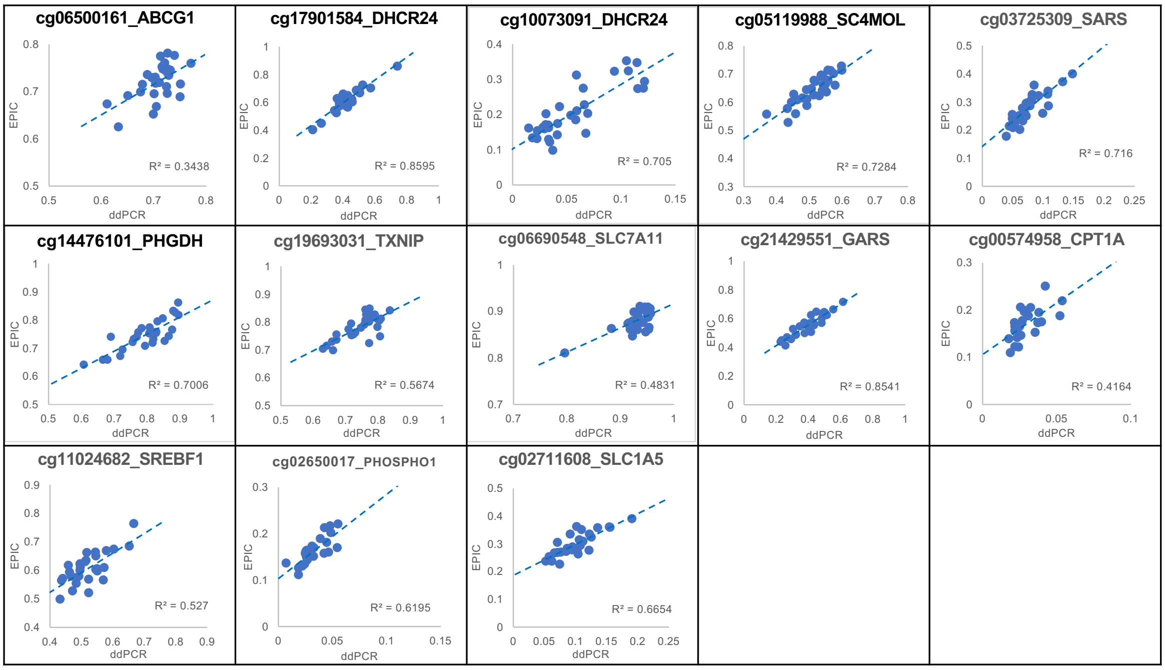

**Fig. S5: Correlation of DNA methylation measurement between ddPCR and EPIC array.**

Scatter plots show a fair correlation between two analytical methods—ddPCR (in the *x*-axis) and EPIC array (in the *y*-axis)—at 13 CpGs examined in this study.

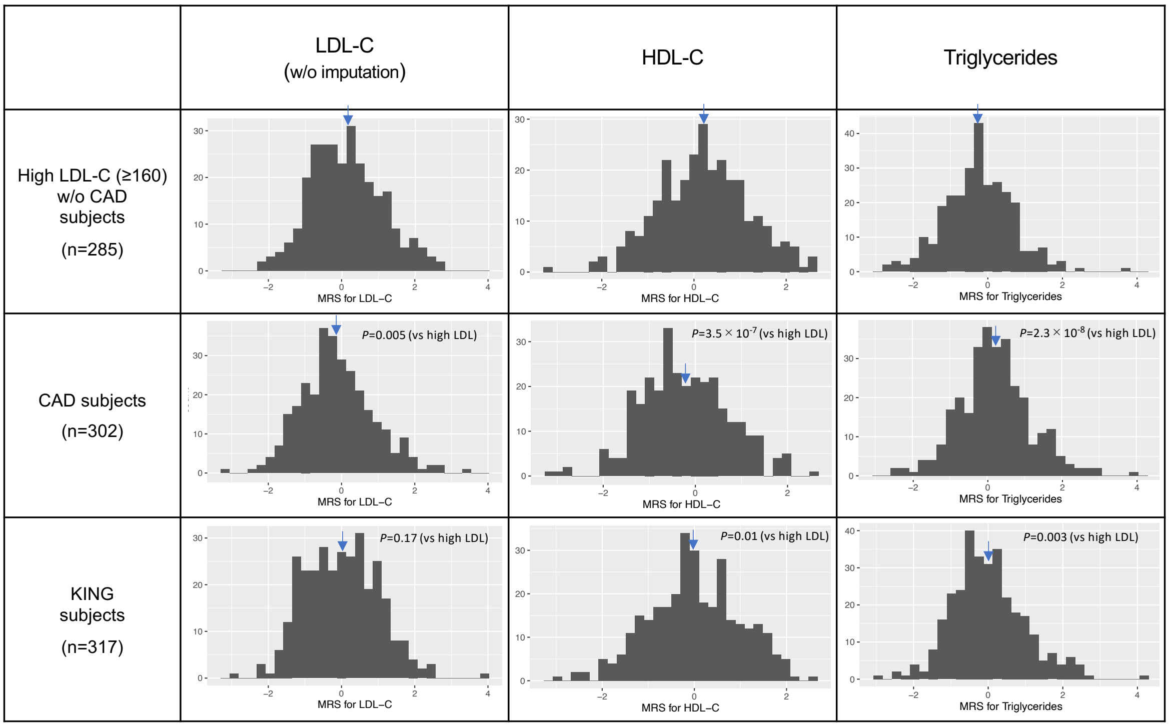

**Fig. S6: Distribution of MRSs for lipid traits calculated with 13 CpGs in high LDL-C, CAD, and general subject groups.**

Histograms of MRSs for lipid traits are shown for two subgroups—high LDL-C (≥160 mg/dl) and CAD individuals—at NCGM and a general subject group (part of the KING study cohort). For those taking lipid-lowering medications (statins or ezetimibe), LDL-C without imputation was used to calculate MRSs in high LDL-C and CAD subgroups to accurately reflect the condition at the time of blood collection. The arrows indicate the mean of lipid MRSs in individual study groups. We made an inter-group comparison of MRSs between the high LDL-C subgroup vs. others.

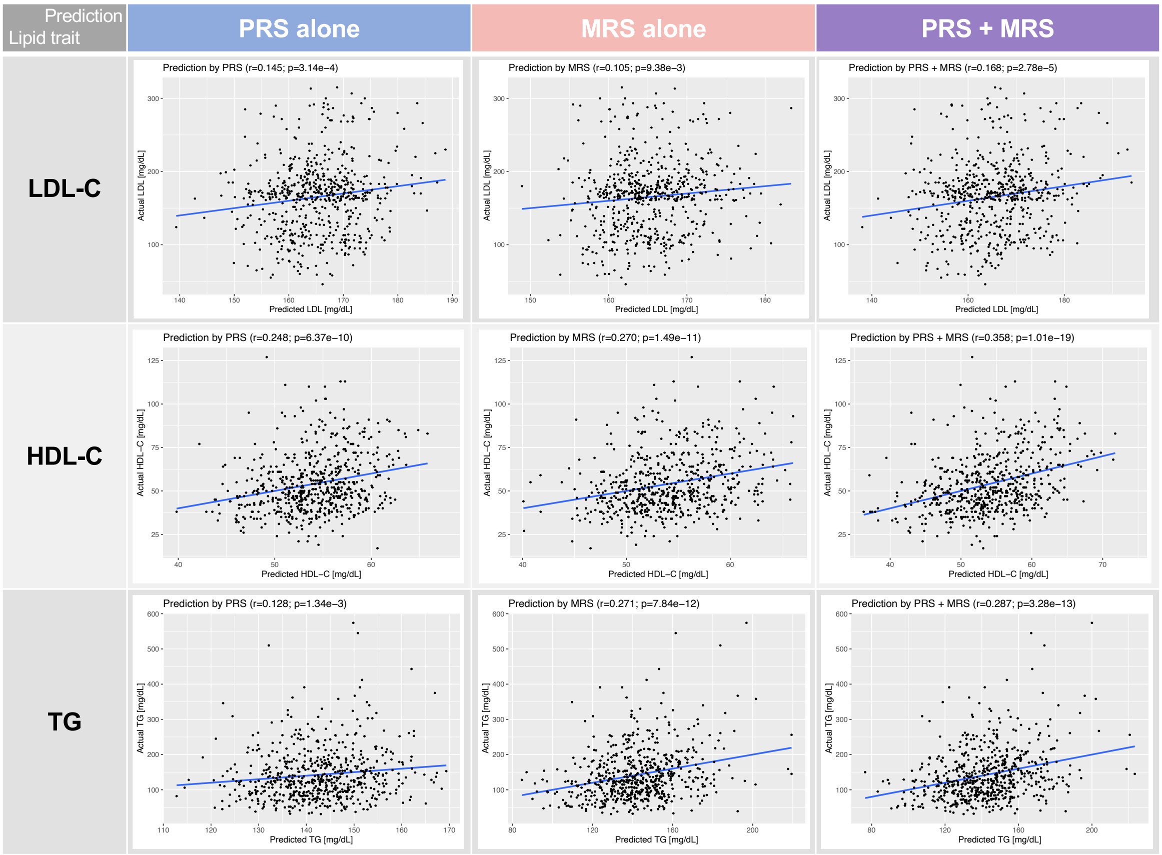

**Fig. S7: Scatter plots demonstrating the correlation between predicted and actual lipid levels using three risk score models.**

Correlation strengths are compared between predicted (in the *x*-axis) and actual (in the *y*-axis) lipid trait values (LDL-C, top panels; HDL-C, middle panels; triglycerides, bottom panels) in each plot using three risk score models: PRS (left column), MRS (middle column), and PRS + MRS (right column). To convert PRS for lipid traits to units of mg/dL, we aligned the PRS with actual lipid measurements in linear regression within the relevant sample, i.e., individuals newly recruited at NCGM (BIO-CVD + NCGM Biobank) for rare variant search besides PRS and MRS calculations (*N* = 653, refer to Fig. 1). For LDL-C, the conversion methods are different between the top panels (of this figure) and Fig. 3D-F, in which an additional conversion formula is used to predict LDL-C for the incorporation of rare variant effects.

Fig.S8

A

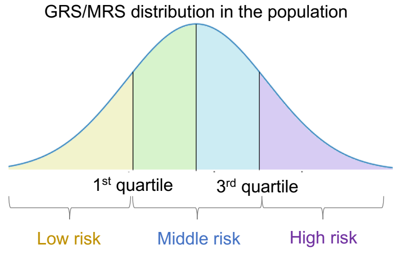

B

| Risk score | PRS (or cGRS) | | | MRS | | |
|---|---|---|---|---|---|---|
| Trait | Low | Middle | High | Low | Middle | High |
| LDL-C | -3.5~-0.5 | -0.5~0.8 | 0.8~4.1 | -5.8~-1.4 | -1.4~1.1 | 1.1~7.2 |
| HDL-C | -8.7~-4.3 | -4.3~-2.2 | -2.2~2.7 | -13.4~-3.4 | -3.4~2.4 | 2.4~11.5 |
| Triglycerides | 1.0~4.1 | 4.1~5.8 | 5.8~9.0 | -54.2~-11.4 | -11.4~10.4 | 10.4~50.9 |

C

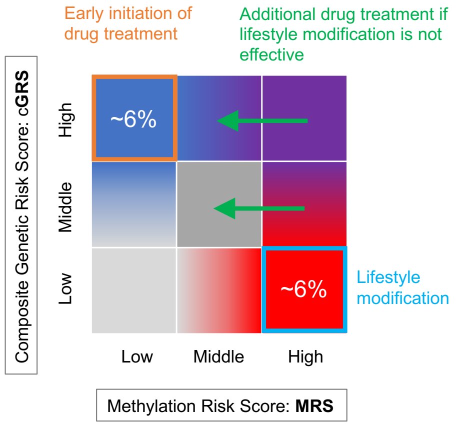

D

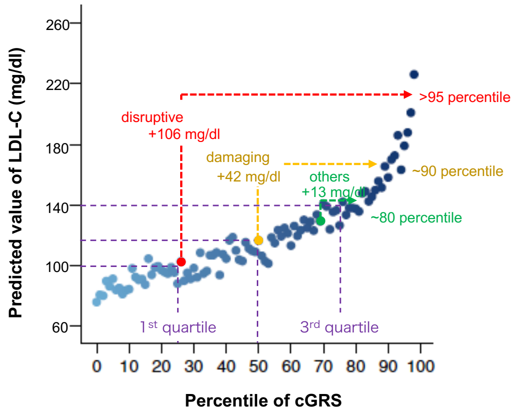

**Fig. S8: Proposed use of dyslipidemia prediction models in clinical practice.**

A policy to prevent or treat dyslipidemia can be determined in clinical practice by using a risk prediction index integrating the composite genetic risk score (or cGRS), which is calculated based on lipid trait-associated SNPs (i.e., PRS) and rare functional variants, and the methylation risk score (MRS). **A**: In terms of the distribution of each risk score in the population, it is broadly classified into a "low-risk group" lower than the first quartile, a "middle-risk group" in the quartile range (first quartile to third quartile), and a "high-risk group" higher than the third quartile. **B**: Specific examples of the GRS and MRS ranges for each lipid trait in the "low-risk," "middle-risk," and "high-risk" groups (based on this study's dataset) are shown in the table. Here, GRSs are based on PRS only, and PRS ranges are based on combined data from three regional populations of Japanese descent (Fig.3A). **C**: Using a matrix table combining two risk scores as a decision support tool, the relative contribution of genetic predisposition and DNA methylation (possible indicators of lifestyle habits) to dyslipidemia can be estimated to standardize and refine policymaking. A higher cGRS reflecting genetic susceptibility tends to indicate a higher need for drug treatment, while a higher MRS reflecting lack of moderation tends to indicate a higher need for lifestyle modification. **D**: For rare functional variant carriers, the predicted value of LDL-C based on the cGRS can be obtained by adding to each person's $PRS_{LDL\text{-}C}$ the estimated changes from non-carriers for each rare variant category (see Methods). If there is a large reference population for which fine quantile information is available, the LDL-C predictions with the correction values for rare functional variants added on top can also be displayed as a percentile.
